# Do polygenic indices capture “direct” effects on child externalizing behavior? Within-family analyses in two longitudinal birth cohorts

**DOI:** 10.1101/2023.05.31.23290802

**Authors:** Peter T. Tanksley, Sarah J. Brislin, Jasmin Wertz, Ronald de Vlaming, Natasia S. Courchesne-Krak, Travis T. Mallard, Laurel L. Raffington, Richard Karlsson Linnér, Philipp Koellinger, Abraham Palmer, Alexandra Sanchez-Roige, Irwin Waldman, Danielle Dick, Terrie E. Moffitt, Avshalom Caspi, K. Paige Harden

## Abstract

Behaviors and disorders characterized by difficulties with self-regulation, such as problematic substance use, antisocial behavior, and symptoms of attention-deficit/hyperactivity disorder (ADHD), incur high costs for individuals, families, and communities. These externalizing behaviors often appear early in the life course and can have far-reaching consequences. Researchers have long been interested in direct measurements of genetic risk for externalizing behaviors, which can be incorporated alongside other known risk factors to improve efforts at early identification and intervention. In a preregistered analysis drawing on data from the Environmental Risk (E-Risk) Longitudinal Twin Study (*N*=862 twins) and the Millennium Cohort Study (MCS; *N*=2,824 parent-child trios), two longitudinal cohorts from the UK, we leveraged molecular genetic data and within-family designs to test for genetic effects on externalizing behavior that are unbiased by the common sources of environmental confounding. Results are consistent with the conclusion that an externalizing polygenic index (PGI) captures causal effects of genetic variants on externalizing problems in children and adolescents, with an effect size that is comparable to those observed for other established risk factors in the research literature on externalizing behavior. Additionally, we find that polygenic associations vary across development (peaking from age 5-10 years), that parental genetics (assortment and parent-specific effects) and family-level covariates affect prediction little, and that sex differences in polygenic prediction are present but only detectable using within-family comparisons. Based on these findings, we believe that the PGI for externalizing behavior is a promising means for studying the development of disruptive behaviors across child development.

**Significance Statement:** Externalizing behaviors/disorders are important but difficult to predict and address. Twin models have suggested that externalizing behaviors are heritable (∼80%), but it has been difficult to measure genetic risk factors directly. Here, we go beyond heritability studies by quantifying genetic liability for externalizing behaviors using a polygenic index (PGI) and employing within-family comparisons to remove sources of environmental confounding typical of such polygenic predictors. In two longitudinal cohorts, we find that the PGI is associated with variation in externalizing behaviors within families, and the effect size is comparable to established risk factors for externalizing behaviors. Our results suggest that genetic variants associated with externalizing behaviors, unlike many other social-science phenotypes, primarily operate through direct genetic pathways.

## Introduction

Behaviors and disorders characterized by difficulties with self-regulation, such as problematic substance use, antisocial behavior, and symptoms of attention-deficit/hyperactivity disorder (ADHD), incur high social costs for individuals, families, and communities. These behaviors, collectively referred to as the externalizing spectrum, often appear early in the life course (by adolescence) and impact a large segment of the youth population (e.g., ∼9% prevalence of ADHD in American youth).^1^ Early manifestations of externalizing behaviors can have far-reaching consequences. For instance, difficulty with self-control assessed during early childhood predicts illness, impecunity, and criminal involvement well into adulthood.^2^ Similarly, people with more conduct problems during development also have more emergency room visits, prescription fills, injury claims, reliance on social welfare, and interaction with the criminal justice system^3^—demonstrating the profound personal and societal costs of externalizing behaviors. Early detection and intervention for externalizing behaviors is, therefore, a public health goal.^2^

Achieving this goal is challenging, however, because human behavior is unpredictable.^4^ Externalizing behaviors are not caused by a single factor, but rather by a complex web of multiple interacting risks. Some of the strongest risk factors for externalizing behavior that have been identified to-date are environmental factors, including exposure to toxins, such as lead^5^ and exposure to maternal smoking *in utero^6^;* social deprivation, including low family socioeconomic status^7^ and neighborhood disadvantage^8^; and relational abuse/trauma, including maltreatment by caregivers^9^ and victimization by peers.^10^ Considered individually, none of these risk factors has an effect size greater than *r* = .2, an effect size that has been characterized as “a medium effect that is of some explanatory and practical use even in the short run” – but is far from a perfect prognosticator.^11^ Identifying additional variables that can contribute to our understanding of who is at elevated risk for the development of externalizing behaviors has the potential to further improve efforts at early detection and intervention.

Behavioral genetic research with twins and adoptees has long hinted that genetic differences contribute to risk for externalizing behavior^12^, but researchers have heretofore been unable to measure that risk directly. Recently, advances in genomic research have raised the possibility of using genetic predictors to supplement efforts at early detection of psychiatric conditions, including those on the externalizing spectrum. Modern genetic predictors are derived from genome-wide association studies (GWAS), wherein the genomes from hundreds of thousands, or even millions, of participants are examined for associations with a common outcome. The results of a GWAS can be aggregated across the whole genome to produce a cumulative measure of genetic liability, or polygenic index (PGI), for the trait under study. Some complex behavioral outcomes, such as educational attainment, are easily quantifiable and have been the subject of large-scale GWAS with sample sizes in the millions.^13^ But the externalizing spectrum is more difficult to quantify owing to the heterogeneity of its expression across the lifespan (e.g., hyperactivity in childhood, substance use in adolescence and adulthood).

Recently, to surmount this challenge, a multivariate GWAS of externalizing behavior, which aggregated genetic information across seven externalizing-related phenotypes, was performed in 1.5 million people of European genetic ancestry.^14^ A PGI constructed from this GWAS was associated with an array of socially important phenotypes in adulthood, including opioid and other substance use, employment histories, and contact with the criminal justice system. Moreover, the externalizing PGI explained ∼10% of the variance in a latent externalizing factor in two independent cohorts, mirroring the variance explained by typical social science variables such as family income^15^ and neighborhood disorder/disadvantage.^16, 17^ These results suggest that an externalizing PGI has potential utility for research aiming to improve the early identification of and intervention on child externalizing problems. However, this PGI analysis was limited because it focused only on late adolescence/adulthood, long after externalizing behaviors have appeared. Here, we build on prior efforts by focusing on early development, in order to more deeply probe the origins of polygenic associations with externalizing behaviors as they appear over the course of development.

Polygenic associations with complex human behavior are complicated to interpret. One interpretation is that they reflect “direct” genetic effects, which are the causal effects of an individual’s own genotype on their own behavior.^18–20^ Note that we place “direct” in quotation marks. This is to indicate that even “direct” genetic effects might be mediated by transactions with the environmental context. For example, a child with genetic predispositions toward disinhibited behavior may evoke harsher punishment from caregivers, which further entrenches the development of conduct problems. Moreover, this negative feedback loop, which leads to an increasing association between child genotype and child behavior over time, might be dampened in families where parents have high levels of social support and material resources, whereas it might be exacerbated in families experiencing stress and deprivation – a gene-environment interaction. Thus, our use of “direct” here refers to genetic effects that originate in one’s own body, but it does not connote the presence or absence of environmental mediators and/or moderators. Put differently, “direct effects incorporate a wide range of causal pathways, some neither simple nor ‘direct’”.^18^

One might assume that PGI associations produce estimates of “direct” genetic effects. However, genotype-phenotype associations may be environmentally confounded because of three processes.^18^ The first, population stratification, arises when groups of people are geographically or socially separated long enough for distinctive patterns of genetic difference to arise through genetic drift. When these groups are compared across social outcomes that also vary between them, it is easy (though wrong) to attribute variation in the outcome to variation in the genetic differences that arose randomly over time.^21^ Second, “indirect” genetic effects can occur because (a) children are genetically similar to others in their environment (e.g., parents, siblings), and (b) many behavioral outcomes are partially influenced by these same others, especially close relations. For example, if a genetic variant increases the likelihood of maternal smoking, and tobacco exposure *in utero* increases the offspring’s risk for conduct problems, then that genetic variant might come to be correlated with conduct problems in a GWAS, but its effect is mediated through an environmental pathway. Third, assortative mating is the process by which people select mates who resemble themselves. Similarity of mate-pairs amplifies the magnitude of indirect genetic effects because it drives up similarity between parents and offspring, as well as between offspring themselves.

Without correction for these sources of confounding, PGI associations will tend to overstate the importance of genetic effects.^18, 22^ Parsing “direct” genetic effects from other sources of genotype-phenotype associations is critical for translating GWAS results into mechanistic knowledge about the biological and social etiology of externalizing behavior, as well as a prudent first step before attempting to incorporate genetic predictors into social and behavioral science. One approach for parsing direct genetic effects from confounding processes is to focus on genetic differences that arise within families.^18, 19^ Within-family designs leverage the random segregation of genotypes that occurs during reproduction, effectively holding population stratification and other sources of environmental confounding constant, and providing more accurate estimates of direct genetic associations.^23, 24^ Put differently, conditional on their parents’ genes, which genes a child inherits is random, such that within-family associations between genotypes and phenotypes are no longer environmentally confounded. Some previous within-family studies have suggested a large role of environmental confounding in PGI associations with other social and behavioral outcomes, such as educational attainment.^25^

In the present study, we estimate within-family PGI associations with youth externalizing behaviors using two analytical designs following our preregistered analytic plan (https://osf.io/nhtw2/). First, the dizygotic twin comparison leverages genetic differences between full siblings, effectively adjusting for genetic effects originating from parental genotypes and the (shared) environmental factors with which parental genotypes may be correlated. Second, the parent-child trio design directly models parental genetic associations with offspring outcomes by including their genotypes in the model, thus making the offspring’s own genotype associations independent of their parents’ genotypes (and independent of environmental factors correlated with parental genotype).^26^ Two previous studies of the externalizing PGI used the parent-child trio design to investigate substance use and disruptive behavior disorders in adolescence to young adulthood, and found evidence that the PGI reflected direct genetic effects rather than environmental confounding.^27–29^ Here, we use two longitudinal cohorts from the UK: the Environmental Risk (E-Risk) Longitudinal Twin Study (*N*=862 same-sex dizygotic twins) and the Millennium Cohort Study (MCS; *N*=2,824 parent-child trios). Our analysis focuses on childhood through adolescence, when externalizing behavior is first and most dynamically expressed. Although externalizing behavior persists throughout adulthood, early development is the period most of interest for intervention.^2^

Our analysis precedes in four steps. First, we contrast population (i.e., between-family) and direct genetic effects on externalizing behavior in our two cohorts across the whole developmental period (i.e., 3-17 years). Second, we test for variation across age groups by dividing development into three separate epochs that characterize major periods of behavioral change: preschool (<5 years), childhood (5-10 years), and adolescence (11-17 years) (see Figure 1 for a depiction of the data collection timeline/developmental epochs observed in each cohort). Third, we examine the sensitivity of PGI population estimates to adjustment for familial contexts, relative to those derived from within-family models. Fourth, and finally, we compare the sensitivity of population and within-family models to sex differences in externalizing behavior across development.

**Figure 1.**
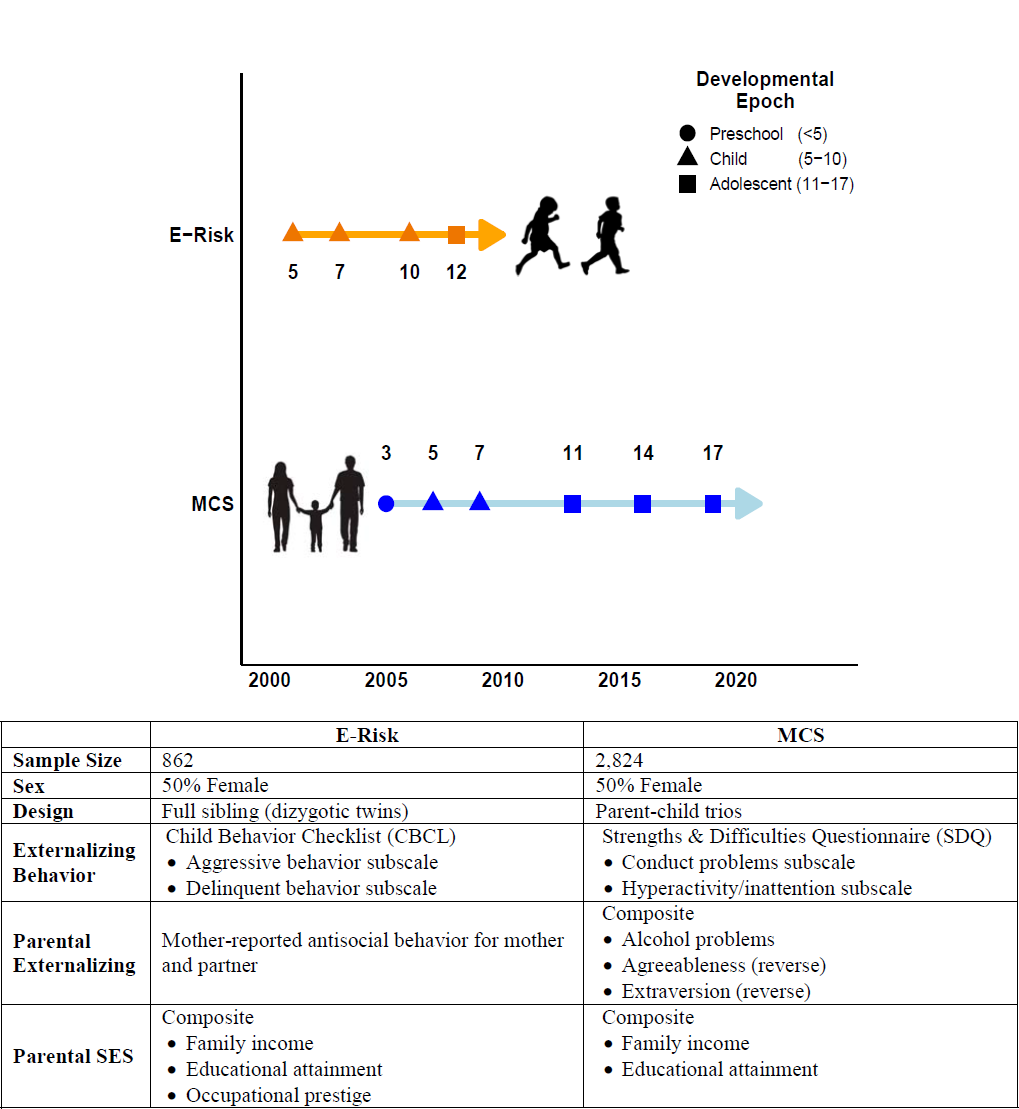
Data collection timeline and key study variables.

## Results

### Young people with higher externalizing PGIs showed more externalizing behavior problems

We constructed genetic and phenotypic measures of externalizing behavior among the European ancestry participants of the E-Risk and MCS cohorts (N=862 and 2,824, respectively) and examined their partial correlation, accounting for age, sex, and the top ten ancestry principal components (to control for residual within-ancestry population stratification). In each cohort, the externalizing PGI (EXT-PGI) was constructed by applying summary statistics from a multivariate GWAS of externalizing behavior in adulthood^14^ to participants’ genetic data (for details, see **Methods** below). In the E-Risk sample, phenotypic externalizing behavior problems in young people was measured as a composite of two subscales from the Child Behavior Checklist (CBCL) assessing delinquent behavior (e.g., swearing, running away) and aggressive behavior (e.g., fighting, threatening). In the MCS, externalizing behavior problems were measured as a composite of two subscales from the Strengths and Difficulties Questionnaire (SDQ) assessing conduct problems (e.g., temper tantrums) and hyperactivity/inattention (e.g., restlessness) (**Supplement** provides further details on measurement). Across cohorts, young people who had higher EXT-PGIs showed more externalizing behavior problems (*r* = .17 for E-Risk; *r* = .2 for MCS; Figure 2A).

**Figure 2.**
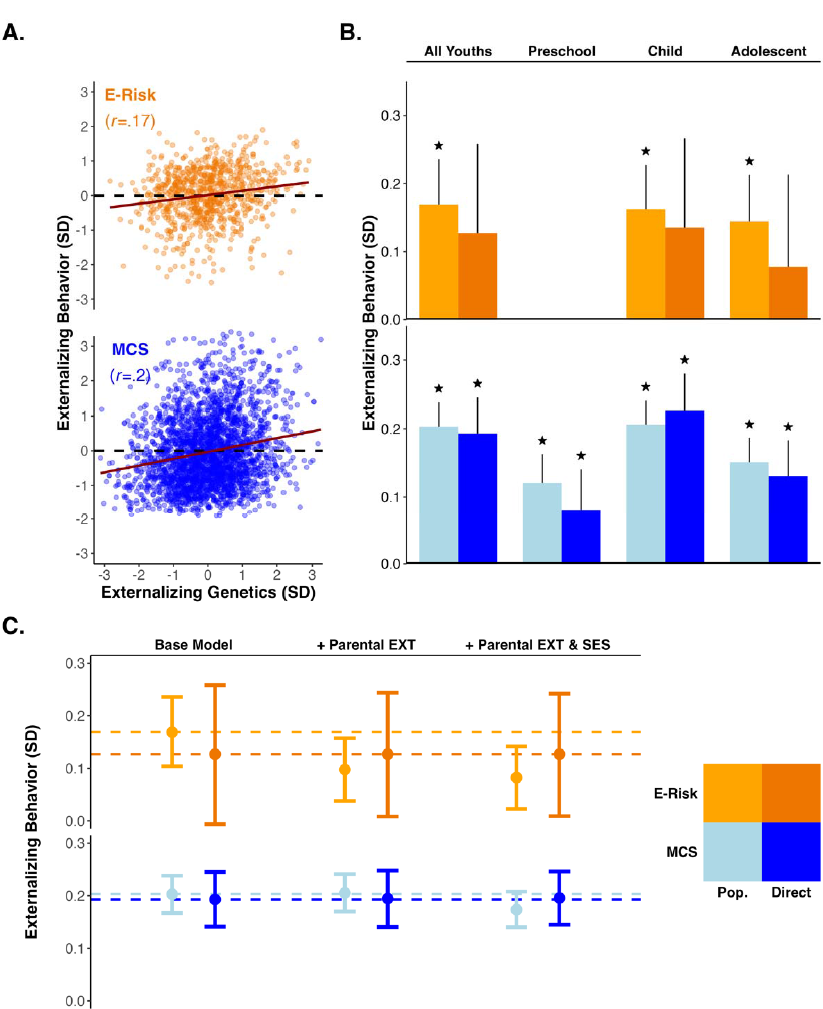
**Panel A**. Scatter plots of the genotypic and phenotypic externalizing measures in the E-Risk (orange) and MCS (blue) cohorts (trendline=slope of correlation). **Panel B**. Bar plot of the population (lighter orange/blue) and direct genetic effects (darker orange/blue) for the E-Risk and MCS cohorts. Columns include bias-adjusted bootstrapped 95% confidence intervals (reps=1000); \**P <* .05 (null: *β*=0). **Panel C**. Point estimates and bias adjusted bootstrapped 95% confidence intervals (reps=1000) across three models of progressive covariate adjustment in the E-Risk and MCS cohorts. The “Base Model” includes no additional covariates, the next model includes parental externalizing behavior, and the final model includes parental externalizing behavior and parental socioeconomic status. The measure of externalizing behavior was derived by pooling observations across all epochs. Dashed horizontal lines provide references for the effect sizes observed in the “Base Model”.

### Young people with higher externalizing PGIs showed more externalizing behavior problems even when comparing within families

We next estimated between- and within-family linear models, pooling all available data within each cohort (Table 1). Across the E-Risk dizygotic twin sample, children with higher values on the EXT-PGI also exhibited more externalizing behavior problems (*β_Population_* = 0.17, 95%CI [0.10, 0.24], *P_FDR_ <* .001). When comparing siblings within the same family to one another, we found the twins with the higher EXT-PGI again had more externalizing behavior problems, on average, than their co-twins (*β_Direct_* = 0.13, 95%CI [-0.002, 0.25], *P_FDR_ =* .077) (Figure 2B, “all youths” model). We note that the estimate from the within-family model was not statistically significant, largely due to the increase in uncertainty typical of sibling fixed effects model.^30^ We observed similar patterns for the children in the MCS cohort, with children who were higher on the EXT-PGI also demonstrating more externalizing problems across development (*β_Population_* = 0.2, 95% CI [0.17, 0.24], *P_FDR_ <* .001). After adjusting for their parents’ genotypes, we found nearly identical associations, which remained statistically significant (*β_Direct_* = 0.19, 95%CI [0.14, 0.25], *P_FDR_ <* .001).

**Table 1.** Population and direct genetic effects of externalizing PGI in the E-Risk and MCS cohorts.

| **E-Risk** |  |  |  |  |  |  |  |  |  |  |  |  |
| --- | --- | --- | --- | --- | --- | --- | --- | --- | --- | --- | --- | --- |
| Effect | **All Youths** |  |  | **Preschool (<5 years)** |  |  | **Child (5-10 years)** |  |  | **Adolescent (11-17 years)** |  |  |
| | $\beta$ | SE | 95% CI | $\beta$ | SE | 95% CI | $\beta$ | SE | 95% CI | $\beta$ | SE | 95% CI |
| Pop. | 0.169 | 0.034*** | [0.104, 0.236] | – | – | – | 0.162 | 0.033*** | [0.098, 0.228] | 0.144 | 0.035*** | [0.077, 0.213] |
| Direct | 0.127 | 0.067 | [-0.006, 0.258] | – | – | – | 0.135 | 0.068 | [0.001, 0.266] | 0.077 | 0.069 | [-0.06, 0.213] |
| Ratio |  | 0.752 |  |  | – |  |  | 0.833 |  |  | 0.535 |  |
| *STD<sub>DIFF</sub>* |  | -0.943 |  |  | – |  |  | -0.581 |  |  | -1.501 |  |
| N |  | 862 |  |  | – |  |  | 862 |  |  | 862 |  |
| **Millennium Cohort Study** |  |  |  |  |  |  |  |  |  |  |  |  |
| Effect | **All Youths** |  |  | **Preschool (<5 years)** |  |  | **Child (5-10 years)** |  |  | **Adolescent (11-17 years)** |  |  |
| | $\beta$ | SE | 95% CI | $\beta$ | SE | 95% CI | $\beta$ | SE | 95% CI | $\beta$ | SE | 95% CI |
| Pop. | 0.203 | 0.018*** | [0.167, 0.238] | 0.121 | 0.021*** | [0.079, 0.162] | 0.206 | 0.018*** | [0.172, 0.241] | 0.151 | 0.018*** | [0.114, 0.186] |
| Direct | 0.193 | 0.026*** | [0.141, 0.245] | 0.08 | 0.030** | [0.022, 0.14] | 0.226 | 0.027*** | [0.173, 0.28] | 0.131 | 0.027*** | [0.078, 0.183] |
| Ratio |  | 0.950 |  |  | 0.667 |  |  | 1.094 |  |  | 0.859 |  |
| *STD<sub>DIFF</sub>* |  | -0.531 |  |  | -1.906 |  |  | 1.093 |  |  | -1.052 |  |
| N |  | 2,824 |  |  | 2,690 |  |  | 2,822 |  |  | 2,813 |  |

\* $p < .05$ , \*\* $p < .01$ , \*\*\* $p < .001$ . $P$ -values are FDR-adjusted. Pop.=population genetic effect of the externalizing PGI on externalizing behavior. Direct=within-family coefficient as estimated by a full-sibling (E-Risk) and parent-child trio (MCS) designs. Ratio=direct genetic effect divided by the population effect. *STD<sub>DIFF</sub>*=standardized difference.

To formally test for attenuation amongst the between-/within-family estimates, we evaluated the standardized difference (*STD_DIFF_*) between *β_Population_* and *β_Direct_* (i.e., a *z*-statistic assumed to be normally distributed; **Supplement** provides materials on the derivation of standard errors).^14^ In the models that pooled data across development, we found that neither the E-Risk (*STD_DIFF_*= −0.94) nor the MCS (*STD_DIFF_* = −0.53) demonstrated statistically significant attenuations in effect size in the within-family models (both two-sided *P* > .05).

These results are consistent with the conclusion that the association between the EXT-PGI and externalizing behavior in young people is primarily explained by direct genetic effects, rather than by other confounding processes, such as population stratification, indirect genetic effects, and assortative mating. However, by pooling data across development, these results could be masking heterogeneity in the effects of the EXT-PGI within specific developmental epochs. We examine this possibility next.

### Genetic associations with externalizing behavior differ across development

We found evidence for a moderate degree of heterogeneity in PGI associations across developmental epochs. We again estimated population and within-family models, but pooled data within discrete developmental epochs instead of across all observations. These epochs were preschool (<5y), childhood (5-10y), and adolescence (11-17y) (see Figure 1 for a depiction of how observations from each cohort fall into each epoch).

Only the MCS provided data on externalizing behaviors within the preschool epoch, which were reported by parents. We found that children younger than five years who had higher values on the EXT-PGI were also reported to have more externalizing behaviors (*β_Population_* = 0.12, 95%CI [0.08, 0.16], *P_FDR_ <* .001). After adjusting for parental genotypes, this association reduced somewhat (*β_Direct_* = 0.08, 95%CI [0.02, 0.14], *P_FDR_ =* .008), although the attenuation was not statistically significant (*STD_DIFF_* = −1.91, two-sided *P* = .057) (Figure 2B and Table 1).

The childhood epoch contained the largest epoch-specific effect sizes for both cohorts. The dizygotic twins in the E-Risk cohort demonstrated the expected pattern of associations, with larger between-family estimates (*β_Population_* = 0.16, 95%CI [0.1, 0.23], *P_FDR_ <* .001) than within-family estimates (*β_Direct_* = 0.13, 95%CI [0.00, 0.26], *P_FDR_ =* .077), but the attenuation remained mild (*STD_DIFF_* = −0.58, two-sided *P* = .561). The MCS cohort exhibited the opposite trend. Children with higher values on the EXT-PGI were predicted to have slightly higher levels of externalizing problems after adjusting for their parents’ genotypes (*β_Direct_* = 0.23, 95%CI [0.17, 0.28], *P_FDR_ <* .001) rather than before (*β_Population_* = 0.21, 95%CI [0.17, 0.24], *P_FDR_ <* .001). However, these differences were again small and non-significant as indicated by a *STD_DIFF_*of 1.09 (two-sided *P* = .275).

We observed a decline in effect sizes for both cohorts in the adolescent epoch. Comparing across all twins in the E-Risk cohort, we found that twins’ EXT-PGI was associated with their externalizing behavior at a level like that of the childhood epoch (*β_Population_* = 0.14, 95%CI [0.08, 0.21], *P_FDR_ <* .001). Unlike the childhood epoch, however, we observed a modest, though statistically nonsignificant, attenuation (*STD_DIFF_* = −1.50, *P* = .133) when comparing twins within families (*β_Direct_* = 0.08, 95%CI [-0.05, 0.21], *P_FDR_ =* .237). In contrast, children in the MCS cohort saw a decline in the association between the EXT-PGI and their externalizing behavior (*β_Population_* = 0.15, 95%CI [0.11, 0.19], *P_FDR_ <* .001) but only a trivial reduction (SDC = −1.05, *P* = 0.293) after accounting for their parents’ genotypes (*β_Direct_* = 0.13, 95%CI [0.08, 0.18], *P_FDR_ =* .237).

Overall, a moderate amount of heterogeneity in effect sizes was uncovered when the models were disaggregated by developmental epochs (*β_Population_’s* ranged from 0.12-0.21), with the largest effect sizes observed in the childhood epoch (5-10y). Despite this, both cohorts demonstrated only mild attenuations in their effect sizes (all attenuations were statistically indistinguishable from zero) when switching to a within-family model, with the largest attenuation occurring in the MCS cohort during the childhood epoch.

### Parental effects do not explain the association between polygenic predictors and externalizing behavior

Next, we investigated two potential sources of bias in our models of population genetic effects: assortative mating and parent-specific genetic effects. (These analyses were not preregistered.) Assortative mating occurs when people are more likely to mate with people who are genotypically and/or phenotypically similar. Assortment increases the variance of a trait in the population and fosters rGE, thus inflating the population genetic effect of a PGI. We assessed assortative mating in the E-Risk cohort by estimating the correlation between dizygotic twins on the EXT-PGI. An elevated correlation with confidence intervals that did not include .5 (i.e., the expected sibling correlation with no assortment) would indicate the presence of assortative mating. The correlation between twins in the E-Risk was *r* = .5 (95%CI [.43, .57]), giving no indication of assortative mating (**Table S1**).

In the MCS, we examined assortative mating related to externalizing genetics by comparing the parents of each trio on their EXT-PGI. Under phenotypic assortment, mate-pair genetics will be independent after adjusting for mate-pair phenotypes.^31^ Thus, mate-pair PGI correlations should be equal to the product of (1) maternal PGI-phenotype correlation, (2) paternal PGI-phenotype correlation, and (3) maternal-paternal phenotype correlation.^13^ The MCS mate-pair correlation on the EXT-PGI was larger than was expected under an assumption of phenotypic assortment (*r_mate-pair_* = .032 vs. *r_Expected_* = .00003); however, the mate-pair correlation was not distinguishable from zero (95% CI [-.004, .067], *P =* .08) leading us to conclude that only negligible assortment on EXT genetics was present in the MCS (**Table S1**).

Next, we compared the relative importance of maternal/paternal genotypes on child externalizing behavior (MCS only). By accounting for one parent at a time (i.e., partially identifying the direct genetic effect), we were able to identify which parental genotypes contributed more to the indirect genetic pathways inflating the population genetic effect. We calculated the percent of attenuation accounted for by adjusting for each parent as:

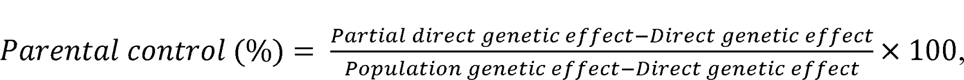

where “partial direct genetic effect” is the estimate for an offspring’s EXT-PGI on their externalizing behavior adjusting for the EXT-PGI of a single parent. When pooling data across epochs, we found that the genotypes of fathers accounted for more of the difference between the population and direct genetic effects (85% vs. 47% for mothers). To contextualize this finding, however, we note that the overall amount of attenuation was very small (*β_Population_*=0.203 vs. *β_Direct_* = 0.198), making even trivial variation in the differences between parents’ estimates appear large. When assessing models within epochs, we found that greatest distance between estimates of parental control was in the childhood epoch, with genotypes of mothers and fathers accounting for 90% and 40%, respectively, of the difference between population and direct genetic effects (**Table S2**).

Overall, these results reinforce the possibility that the EXT-PGI may be driven primarily by direct genetic effects, as there is little evidence for assortment and little difference in effect size between fully and partially adjusted genetic effects.

### Genetic associations with externalizing cannot be accounted for by parental socioeconomic status or parental externalizing behaviors

We examined the robustness of our results by considering family-level phenotypes that are known to be associated with the intergenerational transmission of externalizing behavior: parental externalizing behavior and family socioeconomic status (SES). Including family-level covariates should attenuate estimates of only the population genetic effect, as the within-family model accounts for shared environmental variance by design. However, based on the minimal degree of attenuation seen when comparing the population to the direct genetic effect, we anticipated that only a small portion of the variation in the population genetic effect should be bound up in measured family-level phenotypes.

For both cohorts, we observed no attenuation in the effect sizes of the direct genetic effects and only a small attenuation of the population effect (e.g., largest difference was 0.08 SDs) when including family-level phenotypes (**Table S3** and Figure 2C).

### Sex differences in genetic associations with externalizing were detected only in within-family models

The expression and timing of externalizing behavior across development is different for boys and girls. It is possible that developmental sex differences in externalizing account for some of the above results. We examined this possibility by testing for moderation of the population and direct genetic effects by the sex of the participants.

We recalculated our measure of externalizing behavior in both cohorts by residualizing for age only and averaging between/within developmental epoch. Next, we re-estimated the previous population and within-family models and included terms for *Sex* and *PGI*×*Sex* (**Table S4**). The E-Risk cohort contains only same-sex twin pairs, meaning that only between-pair sex differences could be estimated, as sex only varied at the family level. Because of this, we report results for the E-Risk cohort in the **Supplement** and focus on results from the MCS here. Linear models were used to test for sex differences in the MCS. Effect coding was used for *Sex* to facilitate interpretation. Thus, the intercepts represent the model grand mean of externalizing behavior across the categories in the model (i.e., male, female) for someone of average PGI. The main effects are interpretable as true main effects and not marginal effects. The interaction terms are directly interpretable as the difference in the association (i.e., slope) between the externalizing PGI and phenotypic externalizing for the effect group (i.e., males). Bias-adjusted 95% confidence intervals were produced using *N*=1000 sex-stratified bootstrapped samples to ensure stable interaction estimates.

Pooling data across epochs or examining epochs individually, the between-family model did not identify any statistically significant interactions with sex (**Table S4**). The within-family models identified statistically significant sex difference in all models, save for the childhood epoch. Across all within-family models, excepting the childhood model, the interaction term was positive (*β*’s ranged from 0.13-0.14; all *P_FDR_*<.05) indicating that boys had stronger associations between the externalizing PGI and externalizing behaviors than girls in the MCS. Interestingly, it was the childhood model that also identified the largest main effect of the externalizing PGI across the within-family models (*β_Direct_* = 0.24, 95%CI [0.19, 0.29], *P_FDR_* < .001). The large main effect of the externalizing PGI during the childhood period may help explain the above finding, wherein the direct effect was larger than the population effect during the childhood epoch, as both boys and girls were experiencing similarly high levels of direct genetic effects.

## Discussion

Externalizing behaviors have serious and long-lasting consequences across many life domains. Given their high heritability, there has been great interest in direct measurements of genetic risk that can be incorporated alongside other known risk factors to improve efforts at early identification and intervention. Following a preregistered analytic plan (https://osf.io/nhtw2/), we drew on data from two longitudinal cohorts from the UK, leveraging molecular genetic data and within-family designs to identify genetic effects on externalizing behavior that are unbiased by the common sources of environmental confounding. Results are consistent with the conclusion that an externalizing PGI captures causal effects of genetic variants on externalizing problems in children and adolescents, with an effect size that is comparable to those observed for other established risk factors in the research literature on externalizing (Figure 3).^5-10, 32–34^ For instance, the effect sizes observed in the present work are similar to those seen for childhood maltreatment^9^ and maternal smoking during pregnancy.^6^

**Figure 3.**
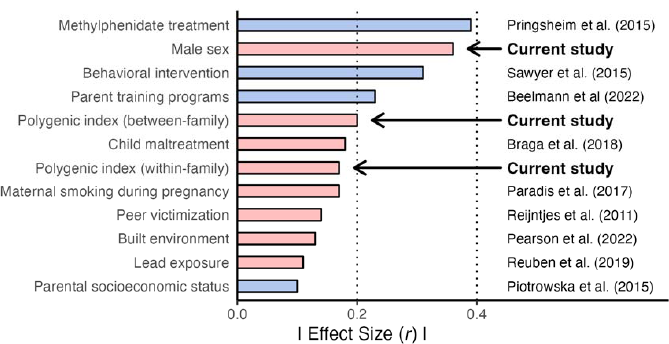
Comparison of effect sizes (interpretable on the correlation scale) of predictors observed in the current study, and those from the literature, on externalizing behavior. Colors represent the polarity of the effect size reported in the original article, with negative values (blue) representing reductions in externalizing behavior and positive values (red) representing increases. Between-/within-family estimates for the externalizing PGI were taken from the MCS models with data pooled across epochs. Estimate for male sex is the main effect of sex from the multiplicative interaction models in the MCS using data pooled across epochs. Note: effect coding was used for sex, making the estimate for sex interpretable as the true main effect, not the marginal effect.

We highlight four key findings. First, we observed only mild attenuation of effect sizes when comparing population and direct effects of the externalizing PGI across cohorts. This reduction is consistent with recent findings where no indirect genetic effects of the externalizing PGI (i.e., residual parental genetic effects after accounting for offspring genotypes) were detected in a Dutch sample of adolescents,^27^ and only weak indirect effects were detected in a sample of American families ascertained for involvement in alcohol treatment programs.^29^

Second, we observed heterogeneity in effect sizes when data were disaggregated by developmental epochs, with the largest effect sizes being observed during the childhood period (5-10yrs) for both cohorts. This result emphasizes the dynamic nature of externalizing behavior across development and highlights the importance of genetic factors early in development. The early peak in effect sizes during childhood and decline in adolescence may be attributable to the increasing heritability of specific forms of externalizing (e.g., alcohol use) above and beyond latent externalizing risk that has been observed later in development.^35, 36^ Despite the variation in mean levels of genetic associations, the differences between population and direct genetic effects were consistently small within epochs, suggesting that the predictive power of the externalizing PGI may vary with developmental epoch but the level of non-direct genetic influence remains low regardless of age.

Third, we found limited impact of statistically adjusting for family-level socioeconomic status and parental externalizing behavior. This result further suggests that the externalizing PGI is not redundant with other, more typically included social science variables. Moreover, we observed only limited evidence for genetic assortment among parents. Overall, the current results suggest only a mild role of the “shared” family-level environment in measured genetic associations with externalizing behavior. This finding accords well with twin studies that estimate that shared, family-wide environmental influences account for ∼20% of the variance in externalizing behaviours.^37^

Fourth, sex differences (i.e., observed in the MCS only) in the association between behavior and the PGI were dependent upon the model used. The between-family models did not detect any sex differences, whereas the within-family models identified sex differences in all but the childhood epoch. We emphasize this last finding as it offers a potential explanation for the general dearth of sex differences observed in polygenic score research.^38^ Although results will vary depending on the nature of the PGI of interest, it may be that the confounds inherent in population genetic associations are sufficient to obfuscate sex differences, and that within-family designs are needed to identify these differences. Except for the childhood epoch (i.e., 5-10yrs), when the PGI had its strongest associations, the current study found that boys demonstrated larger associations between their genetic liability for and expression of externalizing behavior.

This study has several limitations. First, we attempted to maximize comparability across epochs by relying on measures of externalizing that were consistently assessed over time; however, we know that the mode of expression of externalizing behavior is highly varied in early life. It is likely that our approach was not able to capture the full scope of externalizing behaviors as they began to be expressed in the study samples (i.e., due to heterotypic continuity). For example, our externalizing measures did not assess substance use, a form of externalizing behavior that only becomes prominent during adolescence.^39^ The current work may be extended in the future by assessing how the polygenic index for externalizing behavior predicts the expanding range of externalizing behaviors as they onset across development.

Another limitation concerned our samples being restricted to only those participants who had complete data on key study variables, as well as having a complete family structure (e.g., both siblings in the E-Risk and offspring with both parents in the MCS) after listwise deletion. These restrictions may have introduced differential attrition between participants who were and were not included in the final analytic samples and leaving open the possibility of selection effects. This concern is less relevant for the E-Risk sample where retention rates were high (93% of the original sample participated in the most recent wave of data collection). In contrast, only 55% of the original MCS cohort participated in the most recent wave of data collection. We compared the analytic sample from the MCS (*N*=2,824) to the participants with partial data from the relevant waves (*N*=5,377) (**Table S5**). Comparisons revealed that the analytic sample was higher on measures of parental externalizing and SES, lower on genotypic and phenotypic externalizing, and these differences were statistically significant (all *P* < .001). However, effect sizes (Cohen’s *d*) of these differences were small (i.e., ranging from 0.29-0.38), except for parental SES, which was a medium sized effect (|*d*|=0.62) (**Figure S2**). Considering these differences, we cannot rule out the possibility that selection may have influenced some of the results reported here.

Third, while converging results from multiple within-family studies increases confidence that the externalizing PGI associations are not driven entirely, or even predominantly, by environmental confounding, within-family PGI associations are not necessarily driven by the specific genetic variants that are included in the calculation of the PGI. Other genetic variants on the same chromosome that are in linkage disequilibrium with the variants included in the PGI could be the causal variants.^40^ (Two genetic variants are said to be in linkage disequilibrium when they are inherited together more frequently than can be accounted for by chance.) Parsing the specific biological and psychosocial mechanisms that account for the aggregate genetic effects we see here remains a considerable scientific challenge.^41^

Fourth, we relied on a PGI that was originally developed in adults and using phenotypes that are rare or nonexistent in pediatric samples (e.g., number of sexual partners, problematic alcohol use). Despite the PGI being developed on some phenotypes that are misaligned for the present samples, the method used for its construction (i.e., genomic structural equation modeling; GenomicSEM^42^) models the latent trait underlying these and other externalizing phenotypes. Thus, we believe that the genetic underpinnings of the latent externalizing are more likely to be conserved across age^43^ (and be applicable across different expressions of externalizing behavior) than the genetic underpinnings of any specific externalizing behavior.^44^ Additionally, the prior behavioral genetic literature suggests that, while genetic factors change in their importance over development, the sources of genetic effects on externalizing are highly stable. That is, new genetic factors do not come “online” throughout development; the same genetic factors merely become more impactful.^43^

Fifth, our analysis relied solely on British families of European genetic ancestry, and our findings are not expected to be generalizable to children with non-European genetic ancestries. Focusing on European-ancestry children was appropriate for the current analysis because the GWAS of externalizing behavior was conducted in European-ancestry individuals and PGIs (particularly those for complex behavioral traits) have low portability across ancestry groups.^45, 46^ For instance, it has been observed that the PGI for externalizing behavior is less predictive for African-than European-ancestry individuals in an American cohort.^28, 29^ Without a PGI that performs comparably in non-European ancestries, application of the current externalizing PGI to other ancestry groups is unwarranted.

Despite these limitations, the availability of a DNA-based measure of genetic risk for externalizing behavior, which predicts childhood-onset externalizing behaviors even when using a rigorous design-based control for environmental confounding, with an effect size comparable to established risk factors such a family socioeconomic status or lead exposure, is a promising new tool for research that strives to understand and intervene on this pressing public health problem.

## Materials and Methods

### Data sources

Data for this study come from two UK-based cohorts. The Environmental Risk (E-Risk) Longitudinal Twin Study is a prospective birth cohort of 2,232 twins (44% dizygotic) born in 1994-1995 in England and Wales. The sample was assessed at ages 5, 7, 10, 12, and 18.^47^ The analytic sample included only dizygotic twin pairs (50% female) with complete data and who self-identified as White British (*N*=862 twins). Zygosity was confirmed with identity-by-descent estimates (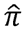) derived from array data.^48^ The Millennium Cohort Study (MCS) is a prospective birth cohort of 18,827 children (18,552 families) born in the United Kingdom at the turn of the new century. The sample was observed at ages 9m, 3, 5, 7, 11, 14, and 17 years of age.^49^ The analytic sample included complete genotyped parent-child trios (children were 50% female) with primarily European ancestry (*N*=2,824). Further details about each cohort are provided in the **Supplement**.

### Polygenic scoring

We computed polygenic indices (PGI) based on the summary statistics from recent GWAS of externalizing behavior.^50^ No significance threshold was applied to select SNPs for inclusion in PGI analyses (i.e., all matched SNPs were included). LD-adjustment was accomplished in the MCS cohort using LDpred2^51^ and in the E-Risk cohort using PRSice-2^52^. Full details about genotyping and PGI construction are provided in the **Supplement**.

### Measures

#### Externalizing

Externalizing behavior was assessed using two behavioral instruments: the Child Behavioral Checklist (CBCL)^53^ in the E-Risk and the Strengths and Difficulties Questionnaire (SDQ)^54^ in the MCS. Following the prior literature, an externalizing measure was derived by combining information from two subscales from each instrument: the aggressive behavior/rule-breaking subscales from the CBCL and the conduct problems and hyperactivity/inattention SDQ subscales. In the E-Risk cohort, the externalizing measure was constructed by averaging each subscale across all reporters (i.e., parent, teacher) for a specific observation then summing the two averages together. The resulting scores were right skewed, so we added a positive constant (1) and log transformed the scale to achieve normality (this process differed from the preregistration). For the MCS, SDQ subscales were averaged across reporters (i.e., parent, teacher, self-report) for each event, and then scores across all events were be entered into a principal component analysis (PCA) and the first PC was extracted. The resulting externalizing scores for both cohorts were then residualized for age, sex, and their interaction, and then averaged across events such that every participant in a cohort has a single age- and sex-independent externalizing score. For developmentally sensitive (i.e., within-epoch) analyses, externalizing scores were residualized and averaged within each developmental period, producing externalizing scores that are age-/sex-independent but specific to developmental periods.

#### Family-level variables

We tested the impact of two family-level mediators through which genetic indirect effects might operate: parental externalizing and parental socioeconomic status (SES).

##### Parental externalizing

In the E-Risk cohort, parental externalizing was measured using antisocial personality disorder symptoms that were reported by the mothers of cohort members for themselves and the twins’ biological fathers when the children were age 5 years. In the MCS cohort, parental externalizing was measured by entering three variables, measured when the children were age 14 years, into a principal components analysis (PCA) and extracting the first PC: an index of alcohol problems (Alcohol Use Disorders Identification Test–Primary Care)^55^ and the agreeableness and conscientiousness subscales (reverse coded) from the “Big Five” personality inventory.^56^ In both cohorts, standardized externalizing composite variables were averaged across parents to produce the final parental externalizing measures.

##### Parental socioeconomic status

In the E-Risk study, we measured parental socioeconomic status (SES) using a standardized composite index of income, education, and social class assessed at age 5 (M=0, SD=1). In the MCS, parental SES was operationalized as a composite of average family income (log) between ages 9m-7yrs and average parental educational attainment (highest earned degree). Both income and educational attainment variables were standardized (mean=0; standard deviation=1) and then averaged across parents to produce the final time-stable parental SES.

#### Covariates

We adjusted for the first ten ancestry PCs to account for population stratification. Both cohorts processed their genotypes in single batches, so no technical covariates were included. We adjust for age, sex, and their interaction; however, these covariates were residualized out of the outcome rather than included in the model alongside other covariates (for a description, see above). Additionally, the E-Risk sample did not include ancestry PCs in analytic models, but instead they were residualized out of the externalizing PGI.

### Analytic plan

#### Genetic effects

In the current study, we identify two distinct genetic effects: the population and direct genetic effects. The population effect includes the direct genetic effects, as well as other sources of confounding (e.g., population stratification, dynastic effects, assortative mating) and is estimated as follows:

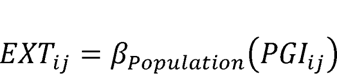

Where phenotypic externalizing, EXT, of individual *i* in family *j* in regressed on their externalizing PGI. Due to the presence of genetic effects other than the direct effects, the *β_Population_* is often inflated beyond the true direct genetic effect (i.e., genetic correlations between the genes of others in the environment and the outcome are absorbed into the population estimate).^18^

Next, we identified the direct genetic effect of the externalizing PGI by leveraging within-family methods developed for the two types of family structures in the current study: full siblings and parent-child trios. Following prior research^57^, we identified the direct genetic effects for full siblings (or dizygotic twins) by estimating a linear model that partitions genetic effects into within- and between-pair genetic effects:

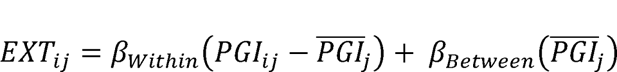

Where *PGI_ij_* is the externalizing PGI value for individual *i* in family *j* and 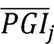 is the family-specific average externalizing PGI value within family *j*. This approach decomposes the PGI association into a within-family component, β*_Within_*, representing the direct genetic effect, versus a between-family component, β*_Between_*, representing the residual genetic effects. Finally, we identified direct genetic effects for parent-child trios using a linear model of the following form:

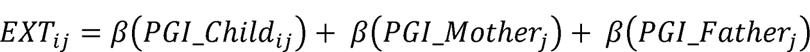

Where *β(PGI_Child_ij_)* captures the direct genetic effect for the child, the residual genetic effects having been adjusted for by the inclusion of both parents’ PGI. All models were estimated as linear models that include the first ten genetic principal components to adjust for population stratification (note: population stratification was adjusted for in the E-Risk by residualizing the externalizing PGI prior to the analysis). Bias-adjusted confidence intervals were estimated using N=1000 bootstrapped samples of each model.

#### Developmental analysis

Using the above methods to identify the population and direct genetic effects, we estimate a series of models to investigate the dynamic nature externalizing behavior across development. The cohorts in the current study observed their participants at different ages throughout development. To maximize comparability across cohorts, we binned observations within three developmentally informed epochs: preschool (<5yrs), childhood (5-10yrs), and adolescence (11-17yrs) (see Figure 1 for a breakdown of the timing of each cohort’s observations relative to developmental epochs).

## Data Availability

Data access is restricted and requires an application to the data owners.

https://cls.ucl.ac.uk/cls-studies/millennium-cohort-study/

https://eriskstudy.com/

## Acknowledgements

**Externalizing Consortium**: We would like to thank the principal investigators and other contributors of the Externalizing Consortium who provided the externalizing summary statistics, as well as editorial suggestions.

**Grant support**: This study was supported by NIH grants 5R01DA050721 (DD) and 5R01HD092548 (KPH), P50DA037844 (AAP), as well as T29KT0526 and T32IR5226 (NIH/NIDA DP1DA054394; ASR).

