## Supplement for "Do polygenic indices capture “direct” effects on child externalizing behavior? Within-family analyses in two longitudinal birth cohorts"

**The role of environmental factors in a genetic predictor of externalizing behavior: Evidence across development in two UK-based cohorts**

**Supplemental Material**

**Sections**

1. **Supplemental Methods**
2. **Supplemental Results**
3. **Supplemental Figures**
4. **Supplemental Tables**
5. **Supplemental References**

**1–Supplemental Methods**

**Cohort descriptions**

E-Risk. We analyze data from the Environmental Risk (E-Risk) Longitudinal Twin Study, which is a longitudinal and nationally representative study that has tracked the development of a birth cohort of 2232 same-sex British twins who were sampled from a birth registry of twins born in England and Wales from 1994 through 1995.^1^ Full details of the E-Risk sample are described elsewhere.^2^ To briefly summarize, the sample was constructed in 1999 to 2000, when 1116 families (93% of those eligible) with same-sex 5-year-old twins participated in home-visit assessments. The full sample was evenly distributed across sex (49% male) and was comprised of 56% monozygotic (MZ; identical) and 44% dizygotic (DZ; fraternal) twin pairs. Families were recruited to represent the United Kingdom population of families with newborns in the 1990s, based on residential location throughout England and Wales and mother’s age. Older mothers having twins via assisted reproduction were under-sampled to avoid an excess of well-educated older mothers, while teenage mothers with twins were over-sampled to ensure enough children growing up in high-risk environments, and to replace teen-mother families lost to the original register due to non-response.

Follow-up home visits were conducted when the participants were aged 7 (98% participation), 10 (96%), 12 (96%), and 18 (93%). The home visits at ages 5, 7, 10, and 12 years included assessments with the participant as well as their mother (or primary caretaker). Each twin participant was assessed by a different interviewer. With parent’s permission, questionnaires were posted to the children’s teachers. Parents gave informed consent and twins gave assent between ages 5 and 12. Twins gave informed consent at age 18 years. Ethical approval for each phase of the study was granted by the Joint South London and Maudsley and the Institute of Psychiatry NHS Ethics Committee. After removing cases with missing values on our key variables and removing MZ twin pairs, our final analytical sample was N = 862 individuals (comprising 431 same-sex DZ twin pairs).

MCS. The Millennium Cohort Study (MCS) is a prospective longitudinal study of 18,827 children (18,552 families) born in the United Kingdom at the turn of the new century.^3^ The MCS is a nationally representative sample with sweeps of data collection when the primary cohort members (the children) were 9m, 3, 5, 7, 11, 14, and 17 years of age. Data was collected on children within the families, as well as their parents/caretakers. Data was collected via surveys administered to children, parents, and teachers, as well as from clinical visits. At age 14, cohort members and their parents were genotyped, making the MCS the only population-based, nationally representative study in the UK containing genetic trios. Following ethical approval for the study from an NHS Research Ethnics Committee (MREC), informed consent is obtained from parents, as well as from the children themselves as they grow up. After removing cases with missing values on our key variables and/or without data for both biological parents, our final sample size was N=2,690-2,824 (sample sizes vary depending on the developmental epoch).

**Genotyping & Imputation**

E-Risk. The Illumina Omni Express 24 BeadChip arrays (twins: Version 1.1, mothers: Version 1.2; Illumina, Hayward, CA) was used to assay common single-nucleotide polymorphism (SNP) variation in the genomes of cohort members and their mothers. We imputed additional SNPs using the IMPUTE2 software (Version 2.3.1; <https://mathgen.stats.ox.ac.uk/impute/impute_v2.html> ^4^) and the 1000 Genomes Phase 3 reference panel.^5^ Imputation was conducted on autosomal SNPs appearing in dbSNP (Version 140; http://www.ncbi.nlm.nih.gov/SNP/)^6^ that were “called” in more than 98% of the samples. Invariant SNPs and SNPs with low minor allele frequency (<1%) were excluded. The E-Risk cohort contains monozygotic twins, who are genetically identical; we therefore empirically measured genotypes of one randomly-selected twin per pair and assigned these data to their monozygotic co-twin. MZ status was confirmed using genotypic data and SNPs from DNA methylation data for subsets of the sample. We directly measured genotypes of both members of dizygotic twin pairs. Prephasing and imputation were conducted using a 50-million-base-pair sliding window. We restricted our analyses to European-descent study participants (90% of E-Risk participants) because allele frequencies, linkage disequilibrium patterns, and environmental moderators of associations may vary across populations.^7^

MCS. The Oragene® 500 DNA self-collection kit was used to collect saliva from cohort members and parents/caretakers when cohort members were age 14. Saliva kits were processed at the Bristol Bioresource Laboratory and a total of 23,336 samples passed initial quality controls (9,259 cohort members, 8,898 mothers and 5,179 fathers). There are 4,533 mother, child, father “trios”. Genetic ancestry was ascertained for each individual who provided saliva samples by projecting MCS data onto the 1000 Genomes data reference panel (phase 3). Based on values from the first five k clusters, the MCS sample was found to be comprised of 86.75% individuals of European ancestry, 10.46% of South Asian ancestry, 2.37% of African ancestry, 0.34% of East Asian ancestry, and 0.07% of Native American ancestry. Genotypes were imputed to the Haplotype Reference Consortium (HRC) reference panel (HRC r1.1; <http://www.haplotypereference-consortium.org>)^8^ using the Michigan Imputation Server (imputationserver.sph.umich.edu).^9^ Phasing was carried out with Eagle.v2.4^10^ and imputed using Minimac.v4.^11, 12^ For the current analysis, only individuals of European ancestry were retained in the analytic sample.

**Polygenic Indices**

We computed polygenic indices (PGI) based on the summary statistics from recent GWAS of externalizing behavior.^13^ Briefly, the externalizing GWAS employed genomic structural equation modeling (genomicSEM^14^) to estimate a latent externalizing factor from seven indicator GWAS, including: attention-deficit/hyperactivity disorder (ADHD)^15^, alcohol dependence^16^, cannabis use^17^, number of sexual partners, age at first sex, and risk tolerance^18^, and smoking.^19^ After estimating the latent externalizing factor from all seven indictor GWAS summary statistics, a multivariate GWAS was then conducted on the latent factor that identified more than 500 independent genome-wide significant hits. No significance threshold was applied to select SNPs for inclusion in PGI analyses (i.e., all matched SNPs were included). Summary statistics were obtained by request through the Externalizing Consortium’s website (externalizing.org).

E-Risk. In the E-Risk cohort, summary statistics for externalizing were adjusted for linkage disequilibrium (LD) using PRSice-2.^20^ The resulting LD-adjusted betas were then applied to the E-Risk sample to calculate an externalizing PGI. To adjust for population stratification, the PGI was then residualized for the first ten genetic ancestry principal components. The PGI was standardized (M=0, SD=1) for each analysis.

MCS. In the MCS cohort, summary statistics for externalizing were adjusted for LD using LDpred-2.^21^ The resulting LD-adjusted betas were then applied to the MCS cohort to calculate an externalizing PGI. Population stratification was adjusted for by including the first ten genetic ancestry principal components in each analytic model of the study. The PGI was standardized (M=0, SD=1) for each analysis.

**Externalizing Behavior**

Externalizing behavior was assessed using two behavioral instruments: the Child Behavioral Checklist (CBCL) in the E-Risk and the Strengths and Difficulties Questionnaire (SDQ) in the MCS.

**E-Risk**. Externalizing behavior was measured as a composite of the delinquent and aggressive behavior subscales from the CBCL^22, 23^ reported by mothers and teachers at ages 5, 7, 10, and 12. This was accomplished by first averaging each subscale across reporters at each event and then summing the average subscale scores together. This resulted in a right skew, so we added a positive constant (1) and log transformed the scale to achieve normality before standardizing (M=0, SD=1) the final measure (this process differed from the preregistration).

**MCS**. Externalizing behavior was measured as a composite of the conduct problems and hyperactivity/inattention subscales of the SDQ^24^ reported by parent at ages 3, 5, 7, 11, 14, 17, by teachers at ages 7 and 17, and cohort members at age 14. This was accomplished by first averaging each subscale across reporters (i.e., parent, teacher, cohort member) and then subscale averages across all events were entered into a principal component analysis (PCA) and the first PC was extracted (M=0, SD=1).

Prior to analysis, the final externalizing scores for both cohorts were residualized for age, sex, and their interaction, and then averaged across events such that every participant in a cohort had a single age- and sex-independent externalizing score (M=0, SD=1). For developmentally sensitive analyses, externalizing scores were residualized and averaged within each developmental epoch, producing externalizing scores that are age-/sex-independent but specific to developmental periods. For instance, E-Risk measure of externalizing included observations from ages 5, 7, and 9 in the childhood epoch and observations from age 12 in the adolescent epoch. For the MCS, the externalizing measure included observations from age 3 for the preschool epoch, ages 5 and 7 for the childhood epoch, and observations from ages 11, 14, and 17 for the adolescent epoch.

**Family-level covariates**

Parental externalizing. In the E-Risk cohort, parental externalizing was measured using antisocial personality disorder symptoms that were reported by the mothers of cohort members for themselves and the twins’ biological fathers at age 5. Symptom counts were right skewed, we thus added a positive constant (1) and performed a log transformation before standardizing the measures (M=0, SD=1). In the MCS cohort, parental externalizing was measured by entering three variables, measured at age 14, into a principal components analysis (PCA): an index of alcohol problems (Alcohol Use Disorders Identification Test–Primary Care) and the agreeableness and conscientiousness subscales (reverse coded) from the “Big Five” personality inventory.^25^ The first PC was extracted (M=0, SD=1), which explaining 40% of the variance. In both cohorts, standardized externalizing composite variables were averaged across parents to produce the final parental externalizing measures.

Parental socioeconomic status. In the E-Risk study, we measured parental socioeconomic status (SES) using a standardized (M=0, SD=1) composite index of income, education, and social class assessed at age 5. In the MCS, parental SES was operationalized as a composite of average family income (log) between ages 9m-7yrs and average parental educational attainment (highest earned degree). Both income and educational attainment variables were standardized (M=0; SD=1) and then averaged across parents to produce the final time-stable parental SES.

**The derivation of standard errors for the difference in OLS coefficients estimated using between- and within-family designs**

Section author: Ronald de Vlaming

**Siblings model**

#### 1.1 Assumed data-generating process

Here, we closely follow derivations by Karlsson Linnér et al. (2021)^13^, found in the Supplementary Information, Section 5.2.6. That is, we assume the following model applies:

$\mathbf{y}=\mathbf{X}\beta+\mathbf{D}\gamma+\varepsilon$,

(1)

where **y** denotes the *N* ×1 outcome vector, **X** the *N* × *K* matrix of explanatory variables with effects in *K* ×1 vector β, ε the *N* ×1 vector of error terms, and **D** the *N* × *F* matrix of family dummies with associated fixed effects in conformable vector γ. For each family *f* = 1,...,*F*, we have data on *n_f_* siblings. Thus, *N* = *n_1_* + *n_2_* +...+ *n_F_* is the total number of observations. Without loss of generality, we assume **X** does not include the intercept—the intercept is perfectly multicollinear with **D**.

#### 1.2 Model omitting the family-specific fixed effects

If the model in Equation 1 holds true, the OLS estimator of β, when omitting **D** and instead only correcting for the intercept, can be written as follows using the Frisch–Waugh–Lovell (FWL) Theorem:

$$\hat{\beta}_{0}=\left( \mathbf{X}_{c}^{\top}\mathbf{X}_{c} \right)^{-1}\mathbf{X}_{c}^{\top}\mathbf{y}_{c}$$

(2)

where **X**_c_ is obtained by mean-centering each column of **X** and **y**_c_ is the mean-centered phenotype vector. That is,

|  | $\mathbf{X}_{c}=\mathbf{CX}, and$ $\mathbf{y}_{c}=\mathbf{Cy}, where$ $\mathbf{C}=\mathbf{I}_{N}-\frac{1}{N}\iota\iota^{\top},$ | (3)  (4)  (5) |
| --- | --- | --- |

where **I***_N_* denotes the *N* × *N* identity matrix and *ι* the *N* × 1 vector of ones. As **C** is a projection matrix, it is idempotent and symmetric (i.e., **CC** = **C** and **C**^⊤^ = **C**). Hence, we have that $\mathbf{X}_{c}^{\top}\mathbf{y}_{c}=\mathbf{X}^{\top}\mathbf{CCy}=\mathbf{X}^{\top}\mathbf{C}^{\top}\mathbf{y}=\mathbf{X}_{c}^{\top}\mathbf{y}$. Hence, we can write $\hat{\beta}_{0}$ as follows:

|  | $\hat{\beta}_{0}=\left( \mathbf{X}_{c}^{\top}\mathbf{X}_{c} \right)^{-1}\mathbf{X}_{c}^{\top}\mathbf{y}$ $=\left( \mathbf{X}_{c}^{\top}\mathbf{X}_{c} \right)^{-1}\mathbf{X}_{c}^{\top}\left( \mathbf{X}\beta+\mathbf{D}\gamma+\varepsilon\right)$ $=\left( \mathbf{X}_{c}^{\top}\mathbf{X}_{c} \right)^{-1}\mathbf{X}_{c}^{\top}\mathbf{X}\beta+\left( \mathbf{X}_{c}^{\top}\mathbf{X}_{c} \right)^{-1}\mathbf{X}_{c}^{\top}\mathbf{D}\gamma+\left( \mathbf{X}_{c}^{\top}\mathbf{X}_{c} \right)^{-1}\mathbf{X}_{c}^{\top}\varepsilon$ | (6)  (7)  (8) |
| --- | --- | --- |

Again using the fact that **C** is idempotent, we have that⊤ $\mathbf{X}_{C}^{\top}\mathbf{X}=\mathbf{X}^{\top}\mathbf{CX}=\mathbf{X}^{\top}\mathbf{CCX}=\mathbf{X}^{\top}\mathbf{C}^{\top}\mathbf{CX}=\mathbf{X}_{C}^{\top}\mathbf{X}_{C}$. Hence,

|  | $\hat{\beta}_{0}=\left( \mathbf{X}_{C}^{\top}\mathbf{X}_{C} \right)^{-1}\mathbf{X}_{C}^{\top}\mathbf{X}_{C}\beta+\left( \mathbf{X}_{C}^{\top}\mathbf{X}_{C} \right)^{-1}\mathbf{X}_{C}^{\top}\mathbf{D}_{\gamma}+\left( \mathbf{X}_{C}^{\top}\mathbf{X}_{C} \right)^{-1}\mathbf{X}_{C}^{\top}\varepsilon$ $=\beta+\left( \mathbf{X}_{C}^{\top}\mathbf{X}_{C} \right)^{-1}\mathbf{X}_{C}^{\top}\mathbf{D}_{\gamma}+\left( \mathbf{X}_{C}^{\top}\mathbf{X}_{C} \right)^{-1}\mathbf{X}_{C}^{\top}\varepsilon.$ | (9)  (10) |
| --- | --- | --- |

Let σ^2^ denote the variance of the error term. Under classical linear model (CLM) assumptions, it now follows that

$$\hat{\beta}_{0}\mathcal{\sim N}\left( \beta+\left( \mathbf{X}_{C}^{\top}\mathbf{X}_{C} \right)^{-1}\mathbf{X}_{C}^{\top}\mathbf{D}_{\gamma},\sigma^{2}\left( \mathbf{X}_{C}^{\top}\mathbf{X}_{C} \right)^{-1} \right).$$

(11)

This distribution of $\hat{\beta}_{0}$ can be interpreted as follows: omission of **D** leads to a bias in $\hat{\beta}_{0}$ in case there are family-specific differences in means of **X** and **y**. Yet, omission of **D** does not affect the *theoretical variance* of $\hat{\beta}_{0}$. On a practical note, however, in a classical application of OLS, $\sigma^{2}$ estimated using the residuals from the regression of **y** on **X** and ι can be considerably larger than the $\sigma^{2}$ estimated from the regression of **y** of **X** and **D**, if the family-specific fixed effects capture a lot of variance in **y**.

#### 1.3 Model accounting for the family-specific fixed effects

Using the FWL Theorem, the OLS estimator of β, in the model in Equation 1, when controlling for **D** can be written as

|  | $\beta_{1}=\left( \mathbf{X}^{\top}\mathbf{MX} \right)^{-1}\mathbf{X}^{\top}\mathbf{My}, where$ $M=\mathbf{I}_{N}-\mathbf{D}\left( \mathbf{D}^{\top}\mathbf{D} \right)^{-1}\mathbf{D}^{\top}$ | (12)  (13) |
| --- | --- | --- |

where **M** is an idempotent, symmetric matrix that projects onto the orthogonal complement of the column space of **D**. Under CLM assumptions, for this estimator we have that

$$\hat{\beta}_{0}\mathcal{\sim N}\left( \beta,\sigma^{2}\left( \mathbf{X}^{\top}\mathbf{MX} \right) \right)$$

(14)

Conceptually, $\hat{\beta}_{0}$ is an estimator of the total association, whereas $\hat{\beta}_{1}$ is an estimator of the within-family (WF) effects.

#### 1.4 Estimated model

In addition to estimating the WF effect, we also wish to quantify the between-family (BF) contribution to the total association. Effectively, we seek to decompose the total association into a WF contribution and BF contribution. We do so by estimating the following model:

$y=\iota a+\mathbf{X}_{\mathrm{WF}}\beta_{\mathrm{WF}}+\mathbf{X}_{\mathrm{BF}}\beta_{\mathrm{BF}}+\eta$,

(15)

where α denotes the intercept, *η* is an *N* × 1 vector of error terms, **X**_1_ is the *N* × *K* matrix of explanatory variables, where each observation of each variable has been adjusted for its family-specific mean for that particular variable, and where **X**_2_ is the *N* × *K* matrix of family-specific means, for each variable and each observation.

Importantly, **X**_WF_ and **X**_BF_ can be written as simple linear algebraic functions of **X** and **D**, *viz*.,

|  | $\mathbf{X}_{\mathrm{WF}}=\mathbf{MX}, and$ $\mathbf{X}_{\mathrm{BF}}=\mathbf{HD}, where$ $\mathbf{H}=\mathbf{D}\left( \mathbf{D}^{\top}\mathbf{D} \right)^{-1}\mathbf{D}^{\top},$ | (16)  (17)  (18) |
| --- | --- | --- |

where **H** is an idempotent, symmetric matrix that projects onto the column space of **D**. Projection matrices **H** and **M** are orthogonal, in the sense that **HM** = **MH** = **0**.

By applying the FWL Theorem and recognizing that **C**ι = **0**, we have that the OLS estimates for β_WF_ and β_BF_ are equivalent to those obtained from applying OLS to the following model:

$$\mathbf{Cy}=\mathbf{C}\mathbf{X}_{\mathrm{WF}}\beta_{\mathrm{WF}}+\mathbf{CX}_{\mathrm{BF}}\beta_{\mathrm{BF}}+\mathbf{C}\eta$$

(19)

By again applying the FWL Theorem, the OLS estimator of β_WF_ when controlling for both **X**_2_ and the intercept can be written as follows:

|  | $\hat{\beta}_{\mathrm{WF}}=\left( \mathbf{X}_{\mathrm{WF}}^{\top}\mathbf{CQC}\mathbf{X}_{\mathrm{WF}} \right)^{-1}\mathbf{X}_{\mathrm{WF}}^{\top}\mathbf{CQCy}, where$ $\mathbf{Q}= \mathbf{I}_{N}-\mathbf{C}\mathbf{X}_{\mathrm{BF}}\left( \mathbf{X}_{\mathrm{BF}}^{\top}\mathbf{C}\mathbf{X}_{\mathrm{BF}} \right)^{-1}\mathbf{X}_{\mathrm{BF}}^{\top}\mathbf{C}.$ | (20)  (21) |
| --- | --- | --- |

Substituting **X**_WF_ and **X**_BF_ by our expressions on the right-hand side of Equations 16–17 allows us to write this WF estimator as follows:

|  | $\hat{\beta}_{\mathrm{WF}}=\left( \mathbf{X}^{\top}\mathbf{MCQCMX} \right)^{-1}\mathbf{X}^{\top}\mathbf{MCQCy}, where$ $\mathbf{Q}= \mathbf{I}_{N}-\mathbf{CHX}\left( \mathbf{X}^{\top}\mathbf{HCHX} \right)^{-1}\mathbf{X}^{\top}\mathbf{HC}.$ | (22)  (23) |
| --- | --- | --- |

Notice here that

|  | $\mathbf{MC}=\left( \mathbf{I}_{N}-\mathbf{D}\left( \mathbf{D}^{\top}\mathbf{D} \right)^{-1}\mathbf{D}^{\top} \right)\left( \mathbf{I}_{N}-\frac{1}{N}\iota\iota^{\top} \right)$ $=\mathbf{I}_{N}-\mathbf{D}\left( \mathbf{D}^{\top}\mathbf{D} \right)^{-1}\mathbf{D}^{\top}-\frac{1}{N}\iota\iota^{\top}+\frac{1}{N}\mathbf{D}\left( \mathbf{D}^{\top}\mathbf{D} \right)^{-1}\mathbf{D}^{\top}\iota\iota^{\top}.$ | (24)  (25) |
| --- | --- | --- |

Here, $\mathbf{D}\left( \mathbf{D}^{\top}\mathbf{D} \right)^{-1}\mathbf{D}^{\top}\iota$ is the projection of the vector of ones (i.e., ι) onto the space spanned by the family-specific dummies, but ι being perfectly multicollinear with **D** means that $\mathbf{D}\left( \mathbf{D}^{\top}\mathbf{D} \right)^{-1}\mathbf{D}^{\top}\iota=\iota$. Hence,

|  | $\mathbf{MC}=\mathbf{I}_{N}-\mathbf{D}\left( \mathbf{D}^{\top}\mathbf{D} \right)^{-1}\mathbf{D}^{\top}-\frac{1}{N}\iota\iota^{\top}+\frac{1}{N}\iota\iota^{\top}$ $=\mathbf{I}_{N}-\mathbf{D}\left( \mathbf{D}^{\top}\mathbf{D} \right)^{-1}\mathbf{D}^{\top}=\mathbf{M}$ | (26)  (27) |
| --- | --- | --- |

The intuition here is that once you ‘partial out’ family-specific fixed effects, using the dummies in **D**, there is nothing to ‘partial out’ for the intercept ι: **M** already sets sample means to zero, by mean-centering within each family.

Therefore, we can rewrite the expression in Equation 22 as follows:

$$\hat{\beta}_{\mathrm{WF}}=\left( \mathbf{X}^{\top}\mathbf{MQMX} \right)^{-1}\mathbf{X}^{\top}\mathbf{MQCy}.$$

(28)

Now, observe that

|  | $\mathbf{MQ}=\mathbf{M}-\mathbf{MCHX}\left( \mathbf{X}^{\top}\mathbf{HCHX} \right)^{-1}\mathbf{X}^{\top}\mathbf{HC}$ $=\mathbf{M}-\mathbf{MHX}\left( \mathbf{X}^{\top}\mathbf{HCHX} \right)^{-1}\mathbf{X}^{\top}\mathbf{HC}$ $=\mathbf{M}-\mathbf{0X}\left( \mathbf{X}^{\top}\mathbf{HCHX} \right)^{-1}\mathbf{X}^{\top}\mathbf{HC}$ $=\mathbf{M}-\mathbf{0}=\mathbf{M}$ | (29)  (30)  (31)  (32) |
| --- | --- | --- |

Thus, the WF estimator can be written as

$$\hat{\beta}_{\mathrm{WF}}=\left( \mathbf{X}^{\top}\mathbf{MX} \right)^{-1}\mathbf{X}^{\top}\mathbf{My}.$$

(33)

This expression is equivalent to $\hat{\beta}_{1}$ in Equation 12. Hence, the OLS estimator of the WF effect is equal to the OLS estimator for β in the model in Equation 1.

Now, let $\hat{\delta}$ denote the difference between $\hat{\beta}_{\mathrm{WF}}$ and the total estimated effect $\hat{\beta}_{0}$. This difference between estimators can be written as follows:

|  | $\hat{\delta}=\hat{\beta}_{\mathrm{WF}}-\hat{\beta}_{0}$ $=-\left( \mathbf{X}_{C}^{\top}\mathbf{X}_{C} \right)^{-1}\mathbf{X}_{C}^{\top}\boldsymbol{D\gamma+}\left( \left( \mathbf{X}^{\top}\mathbf{MX} \right)^{-1}\mathbf{X}^{\top}\mathbf{M}-\left( \mathbf{X}_{C}^{\top}\mathbf{X}_{C} \right)^{-1}\mathbf{X}_{C}^{\top} \right)\varepsilon.$ | (34)  (35) |
| --- | --- | --- |

Consequently, under CLM assumptions, we have that

$$\hat{\delta}\mathcal{\sim N}\left( -\left( \mathbf{X}_{C}^{\top}\mathbf{X}_{C} \right)^{-1}\mathbf{X}_{C}^{\top}\mathbf{D}\gamma,\sigma^{2}\left( \left( \mathbf{X}^{\top}\mathbf{MX} \right)^{-1}\mathbf{X}^{\top}\mathbf{M}-\left( \mathbf{X}_{C}^{\top}\mathbf{X}_{C} \right)^{-1}\mathbf{X}_{C}^{\top} \right)\left( \left( \mathbf{X}^{\top}\mathbf{MX} \right)^{-1}\mathbf{X}^{\top}\mathbf{M}-\left( \mathbf{X}_{C}^{\top}\mathbf{X}_{C} \right)^{-1}\mathbf{X}_{C}^{\top} \right)^{\top} \right).$$

As shown by Karlsson Linnér et al. (2021), the variance matrix of this distribution can be simplified, yielding

$$\hat{\delta}\mathcal{\sim N}\left( -\left( \mathbf{X}_{C}^{\top}\mathbf{X}_{C} \right)^{-1}\mathbf{X}_{C}^{\top}\mathbf{D}\gamma,\sigma^{2}\left( \left( \mathbf{X}^{\top}\mathbf{MX} \right)^{-1}-\left( \mathbf{X}_{C}^{\top}\mathbf{X}_{C} \right)^{-1} \right) \right).$$

(36)

This simplification even holds in our slightly more general formulation, where both the expectation and the second term in the variance matrix involve $\mathbf{X}_{C}^{\top}$ instead of **X**, seen in the work by Karlsson Linnér et al. (2021).

#### 1.5 Remark on estimating the error variance

Although the $\hat{\beta}_{\mathrm{WF}}=\hat{\beta}_{1}$, the residuals from the model in Equation 1 and the model in Equation 15 are not necessarily the same. The simple reason is that space spanned by ι, **X**_WF_, and **X**_BF_ jointly is actually a subspace of the space spanned by **X** and **D**. To put it somewhat colloquially, **X** and **D** combined can explain a larger chunk of **y** than ι, **X**_WF_, and **X**_BF_ combined. Hence, we have that $\hat{\eta}^{\top}\hat{\eta}\geq\hat{\varepsilon}^{\top}\hat{\varepsilon}$, where $\hat{\eta}$ denote the OLS residuals from the model in Equation 15 and $\hat{\varepsilon}$ the OLS residuals from the model in Equation 1.

The Python script in Code Listing (see below) highlights the equivalence of the OLS estimators as well as the difference in the residuals, and the resulting difference in the estimated error variance. In the simulation presented in this code, we have *K* = 2 regressors. Figures 1 and 2 shows the estimated coefficients for the two regressors according to $\hat{\beta}_{1}$, and $\hat{\beta}_{\mathrm{WF}}$ across 1000 runs, illustrating their equivalence. Figure 3 shows the estimated error variance based on OLS estimates for the model in Equation 1 and that in Equation 15 across the same set of runs. In this specific simulation design, the estimated error variance based on the former model is consistently lower than that of the latter. Moreover, the estimates of error variance are imperfectly correlated across the two models. Hence, this difference is not just a scaling issue due to different degrees of freedom in the two models (i.e., *N* − (*K* + *F*) for the former and *N* − (2*K* + 1) for the latter).

| **Estimator** | **Estimator intends to capture** | **Residuals for estimating** $\boldsymbol{\sigma}^{\boldsymbol{2}}$ | **Degrees of freedom** ${\hat{\boldsymbol{\sigma}}}^{\boldsymbol{2}}$ |
| --- | --- | --- | --- |
| $\hat{\beta}_{0}=\left( \mathbf{X}_{c}^{\top}\mathbf{X}_{c} \right)^{-1}\mathbf{X}_{c}^{\top}\mathbf{y}_{c}$ | Total association | $e_{0}=\mathbf{y}_{C}-\mathbf{X}_{C}\hat{\beta}_{0}$ | $N-\left( K+1 \right)$ |
| $\beta_{1}=\left( \mathbf{X}^{\top}\mathbf{MX} \right)^{-1}\mathbf{X}^{\top}\mathbf{My}$ | Within-family association | $e_{1}=\mathbf{My}-\mathbf{MX}\hat{\beta}_{1}$ | $N-\left( K+F \right)$ |
| $\hat{\beta}_{\mathrm{WF}}=\left( \mathbf{X}^{\top}\mathbf{MCQCMX} \right)^{-1}\mathbf{X}^{\top}\mathbf{MCQCy}=\hat{\beta}_{1}$ | Within-family association | $e_{\mathrm{WF}}=\mathbf{QCy}-\mathbf{QCMX}\hat{\beta}_{1}$ | $N-\left( 2K+1 \right)$ |
| $\hat{\delta}=\hat{\beta}_{\mathrm{WF}}-\hat{\beta}_{0}$ | Attenuation due to removing between-family variation | $e_{\delta}=e_{1}=\mathbf{My}-\mathbf{MX}\hat{\beta}_{1}$ | $N-\left( K+F \right)$ |
| Table 1: Overview of estimators for the siblings model for F families. We have data on *n_f_* siblings for family *f* = 1,...,*F*. We have *N* = *n*_1_ + ... + *n_F_* observations in total. For each observation, we have data on *K* regressors of interest (excluding the intercept), such as polygenic indices (PGIs). **X** denotes the *N* × *K* matrix of regressors and **D** the *N* × *F* matrix of family-specific dummies. Further definitions: $\mathbf{H}=\mathbf{D}\left( \mathbf{D}^{\top}\mathbf{D} \right)^{-1}\mathbf{D}^{\top}$, $\mathbf{M}=\mathbf{I}_{N}-\mathbf{H}$, $\mathbf{C}=\mathbf{I}_{N}-\frac{1}{N}\iota\iota$, $\mathbf{Q}=\mathbf{I}_{N}-\mathbf{CHX}\left( \mathbf{X}^{\top}\mathbf{HCHX} \right)^{-1}\mathbf{X}^{\top}\mathbf{HC}$, $\mathbf{X}_{C}=\mathbf{CX}$, and $\mathbf{y}_{c}=\mathbf{Cy}$. | | | |

Since our focus lies on the behaviour of $\hat{\delta}$ when we assume the model in Equation 1 holds true, it seems fairest to use residuals from that model to estimate the error variance, $\sigma^{2}$. That is, we calculate

|  | $\hat{\sigma}_{1}^{2}=\frac{1}{N-\left( K+F \right)}e_{1}^{\top}e_{1}, where$ $e_{1}=\mathbf{My}-\mathbf{MX}\hat{\beta}_{1}.$ | (37)  (38) |
| --- | --- | --- |

This estimate (i.e., $\hat{\sigma}_{1}^{2}$) can then be used to set the sampling variance of matrix of $\hat{\delta}$ as follows:

$$\hat{\mathrm{Var}}\left( \hat{\delta} \right)=\hat{\sigma}_{1}^{2}\left( \left( \mathbf{X}^{\top}\mathbf{MX} \right)^{-1}-\left( \mathbf{X}_{c}^{\top}\mathbf{X}_{c} \right)^{-1} \right)$$

(39)

At the same time, to ensure standard errors for $\hat{\beta}_{0}$, $\hat{\beta}_{1}$, and $\hat{\beta}_{\mathrm{WF}}$align will standard OLS output, the error variance there will be estimated directly from the estimated model, rather than residuals from the model that we assume to be the truth.

An overview of estimators and the residuals used to calculate error variance for the siblings model is shown in Table 1.

### 2 Trios model

#### 2.1 Assumed data-generating process

Here, we again assume to have data from *F* different families. However, in each family we observe outcome data for only one child. That is, *n_f_* = 1 for *f* = 1,...,*F*. Consequently, *N* = *F*. In this case, the corresponding matrix of family-specific dummies **D** is *N* × *N* and such that **H** = **I***_N_* and **M** = **0**. In other words, controlling for **D** annihilates all variation y and regressors. Therefore, we impose a different data-generating process (DGP) here. Specifically, we assume

|  | $\mathbf{y}=\mathbf{Z}\gamma+\mathbf{P}\pi+\mathbf{x}\beta+\varepsilon, where$ $\mathbf{P}=\left( \mathbf{x}_{M} \mathbf{x}_{F} \right), and$ $\pi=\left( \begin{matrix} \pi_{M} \\ \pi_{F} \end{matrix} \right),$ | (40)  (41)  (42) |
| --- | --- | --- |

where **x** denotes the *N* ×1 vector of PGI values for the set of individuals under consideration and **x**_M_ (resp. **x**_F_) denotes the *N* × 1 vector of maternal (paternal) PGI values, with effect π_M_ (π_F_). In addition, **Z** is a *N* × *M* matrix of control variables that includes the intercept, with *M* × 1 vector of effects γ.

#### 2.2 Estimated models

Let **R** denote the projection matrix that ‘partials out’ the effects of **Z**. That is,

$$\mathbf{R}=\mathbf{I}_{N}-\mathbf{Z}\left( \mathbf{Z}^{\top}\mathbf{Z} \right)^{-1}\mathbf{Z}^{\top}.$$

(43)

Hence, under the DGP in Equation 40, we have that

$$\mathbf{Ry}=\mathbf{RP}\pi+\mathbf{Rx}\beta+\mathbf{R}\varepsilon.$$

(44)

In addition, let **T** denote the projection matrix that ‘partials out’ the remaining effects of **RP**. That is,

$$\mathbf{T}=\mathbf{I}_{N}-\mathbf{RP}\left( \mathbf{P}^{\top}\mathbf{RP} \right)^{-1}\mathbf{P}^{\top}\mathbf{R}.$$

(45)

Hence, under the DGP in Equation 40, we have that

$$\mathbf{TRy}=\mathbf{TRx}\beta+\mathbf{TR}\varepsilon.$$

(46)

Consequently, the OLS estimates for $\beta$ for the model in Equation 40 can be written as follows:

$$\hat{\beta}=\left( \mathbf{x}^{\top}\mathbf{RTRx} \right)^{-1}\mathbf{x}^{\top}\mathbf{RTRy}.$$

(47)

This expression is basically the result of applying the FWL Theorem twice. This OLS estimator $\hat{\beta}$ can be rearranged as

|  | $\hat{\beta}=\left( \mathbf{x}^{\top}\mathbf{RTRx} \right)^{-1}\mathbf{x}^{\top}\mathbf{R}\left( \mathbf{TRx}\beta+\mathbf{TR}\varepsilon\right)$ $=\left( \mathbf{x}^{\top}\mathbf{RTRx} \right)^{-1}\mathbf{x}^{\top}\mathbf{RTRx}\beta+\left( \mathbf{x}^{\top}\mathbf{RTRx} \right)^{-1}\mathbf{x}^{\top}\mathbf{RTR}\varepsilon$ $= \beta+ \left( \mathbf{x}^{\top}\mathbf{RTRx} \right)^{-1}\mathbf{x}^{\top}\mathbf{RTR}\varepsilon.$ | (48)  (49)  (50) |
| --- | --- | --- |

Hence, under the CLM assumptions, we have that

$$\hat{\beta}\mathcal{\sim N}\left( \beta,\sigma^{2}\left( \mathbf{x}^{\top}\mathbf{RTRx} \right)^{-1} \right).$$

(51)

In addition, we also consider a model in which **P** is omitted, *viz*.,

$$\mathbf{y}={\mathbf{Z}\gamma}_{0}+\mathbf{x}\beta_{0}+\eta,$$

(52)

Where $\eta$ denotes the error term. The OLS estimator of $\beta_{0}$ in this model can be written as:

$$\hat{\beta}_{0}=\left( \mathbf{x}^{\top}\mathbf{Rx} \right)^{-1}\mathbf{x}^{\top}\mathbf{Ry}.$$

(53)

Assuming the data-generating process in Equation 40 holds true, the latter OLS estimator can be written as

|  | $\hat{\beta}_{0}=\left( \mathbf{x}^{\top}\mathbf{Rx} \right)^{-1}\mathbf{x}^{\top}\mathbf{R}\left( \mathbf{TRx}\beta+\mathbf{TR}\varepsilon\right)$ $=\left( \mathbf{x}^{\top}\mathbf{Rx} \right)^{-1}\mathbf{x}^{\top}\mathbf{RP}\pi+\left( \mathbf{x}^{\top}\mathbf{Rx} \right)^{-1}\mathbf{x}^{\top}\mathbf{Rx}\beta+\left( \mathbf{x}^{\top}\mathbf{Rx} \right)^{-1}\mathbf{x}^{\top}\mathbf{R}\varepsilon$ $=\beta+\left( \mathbf{x}^{\top}\mathbf{Rx} \right)^{-1}\mathbf{x}^{\top}\mathbf{RP}\pi+\left( \mathbf{x}^{\top}\mathbf{Rx} \right)^{-1}\mathbf{x}^{\top}\mathbf{R}\varepsilon.$ | (54)  (55)  (56) |
| --- | --- | --- |

Hence, again under the CLM assumptions, we have that

$$\hat{\beta}_{0}\sim\mathcal{N}\left( \beta+\left( \mathbf{x}^{\top}\mathbf{Rx} \right)^{-1}\mathbf{x}^{\top}\mathbf{RP}\pi,\sigma^{2}\left( \mathbf{x}^{\top}\mathbf{Rx} \right)^{-1} \right).$$

(57)

Now, let $\hat{\delta}$ denote the difference between $\hat{\beta}$ and $\hat{\beta}_{0}$. This difference between estimators can be written as follows:

| $\hat{\delta}=\hat{\beta}-\hat{\beta}_{0}$ $=-\left( \mathbf{x}^{\top}\mathbf{Rx} \right)^{-1}\mathbf{x}^{\top}\mathbf{RP}\pi+\left( \left( \mathbf{x}^{\top}\mathbf{RTRx} \right)^{-1}\mathbf{x}^{\boldsymbol{\top}}\mathbf{RTR}-\left( \mathbf{x}^{\top}\mathbf{RTRx} \right)^{-1}\mathbf{x}^{\top}\mathbf{R} \right)\varepsilon.$ | (58)  (59) |
| --- | --- |

Analogous to the derivations by Karlsson Linnér et al. (2021), this implies

$$\hat{\delta}\mathcal{\sim N}\left( -\left( \mathbf{x}^{\top}\mathbf{Rx} \right)^{-1}\mathbf{x}^{\top}\mathbf{RP}\pi,\sigma^{2}\left( \left( \mathbf{x}^{\top}\mathbf{RTRx} \right)^{-1}-\left( \mathbf{x}^{\top}\mathbf{Rx} \right)^{-1} \right) \right).$$

(60)

The regression residuals from the model in Equation 40 can be readily used to estimate the error variance σ^2^. Plugging this estimator into the theoretical variance of $\hat{\delta}$ we obtain the sampling variance:

$$\hat{\mathrm{Var}}\left( \hat{\delta} \right)=\hat{\sigma}^{2}\left( \left( \mathbf{x}^{\top}\mathbf{RTRx} \right)^{-1}-\left( \mathbf{x}^{\top}\mathbf{Rx} \right)^{-1} \right).$$

(61)

**Code Listing 1: Simulation highlighting the equivalence of WF estimators, and the difference in residual variance (sibling design).**

| 1  2  3  4  5  6  7  8  9  10  11  12  13  14  15  16  17  18  19  20  21  22  23  24  25  26  27  28  29  30  31  32  33  34  35  36  37  38  39  40  41  42  43  44  45  46  47  48  49  50  51  52  53  54  55  56  57  58  59  60  61  62  63  64  65  66  67  68  69  70  71  72  73  74  75  76  77  78 | **import** **numpy** **as** **np**  **import** **matplotlib.pyplot** **as** **plt**  **import** **math**  *# set seed for random-number generator*  rng=np.random.default_rng(1238431)  *# set number of families, siblings per family, sample size, regressors, runs*  f=500  c=2  n=f*c  k=2  RUNS=1000  *# set family dummies, and projection matrices based thereon*  D=np.kron(np.eye(f),np.ones((c,1)))  H=D@np.linalg.inv(D.T@D)@D.T  M=np.eye(n)-H  *# initialise matrix to track OLS estimators across runs*  *# 1) WF estimators when controlling for family-specific fixed effects*  B1=np.zeros((k,RUNS))  *# 2) WF estimators from decomposing regressors into BF and WF part*  BWF=np.zeros((k,RUNS))  *# initialise vectors to track residual variance estimates across runs*  V1=np.zeros((RUNS))  V2=np.zeros((RUNS))  *# for each run*  **for** r **in** range(RUNS):  *# set mean per family per regressor*  muX=rng.normal(size=(f,k))  *# set regressor per individual, taking family-specific mean into account*  X=D@muX+rng.normal(size=(n,k))  *# draw family-specific effects*  gamma=rng.normal(size=(f,1))  *# draw true regressor effects*  beta=rng.normal(size=(k,1))  *# draw error term*  eps=rng.normal(size=(n,1))  *# calculate outcome*  y=X@beta+D@gamma+eps  *# get WF estimates when controlling for family-specific fixed effects*  b=np.linalg.inv(X.T@M@X)@(X.T@M@y)  B1[:,r]=b.ravel()  *# calculate residuals and estimate residual variance*  e=(M@y)-((M@X)@b)  V1[r]=((e**2).sum())/(n-(k+f))  *# decompose X into BF and WF part, and use those parts + intercept*  Z=np.hstack((M@X,H@X,np.ones((n,1))))  *# get WF estimates from decomposing regressors into BF and WF part*  b=np.linalg.inv(Z.T@Z)@(Z.T@y)  BWF[:,r]=b[0:k].ravel()  *# calculate residuals and estimate residual variance*  e=y-Z@b  V2[r]=((e**2).sum())/(n-(2*k+1))  **for** j **in** range(k):  fig=plt.figure()  ax=fig.add_subplot(111)  plt.scatter(B1[j,:],BWF[j,:],s=3)  plt.xlim((math.floor(min((min(B1[j,:]),min(BWF[j,:])))),\  math.ceil(max((max(B1[j,:]),max(BWF[j,:]))))))  plt.ylim((math.floor(min((min(B1[j,:]),min(BWF[j,:])))),\  math.ceil(max((max(B1[j,:]),max(BWF[j,:]))))))  ax.set_aspect('equal')  plt.xlabel("Controlling for family-specific fixed effects")  plt.ylabel("Decomposing regressors into BF and WF part")  plt.title("WF estimator for Regressor "+str(j+1)+" across "+str(RUNS)+" runs")  plt.grid()  plt.savefig("beta"+str(j+1)+".pdf")  fig=plt.figure()  ax=fig.add_subplot(111)  plt.scatter(V1,V2,s=3)  plt.xlim((0.9*(min((min(V1),min(V2)))),\  1.1*(max((max(V1),max(V2))))))  plt.ylim((0.9*(min((min(V1),min(V2)))),\  1.1*(max((max(V1),max(V2))))))  ax.set_aspect('equal')  plt.xlabel("Controlling for family-specific fixed effects")  plt.ylabel("Decomposing regressors into BF and WF part")  plt.title("Estimated error variance across "+str(RUNS)+" runs")  plt.grid()  plt.savefig("var.pdf") |
| --- | --- |

| 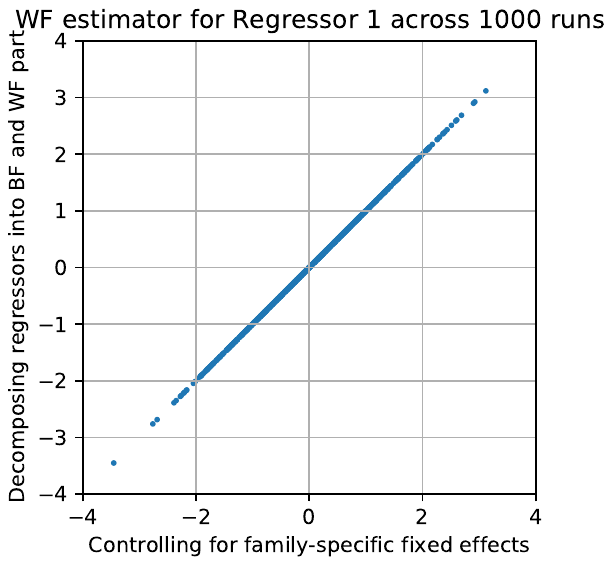 | **Code listing figure 1.** Estimated coefficients for the first regressor according to $\hat{\beta}_{1}$ (x-axis) and $\hat{\beta}_{WF}$ (y-axis), across 1000 runs in the simulation in **Code Listing 1.** |
| --- | --- |
| 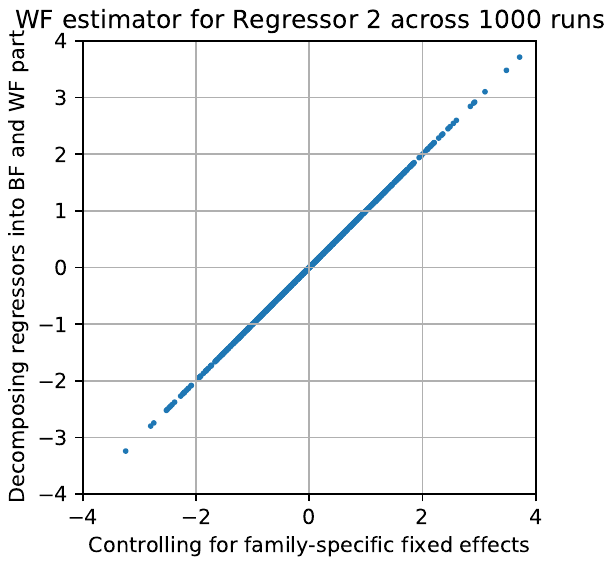 | **Code listing figure 2**. Estimated coefficients for the second regressor according to $\hat{\beta}_{1}$ (x-axis) and $\hat{\beta}_{WF}$ (y-axis), across 1000 runs in the simulation in **Code Listing 1**. |
| 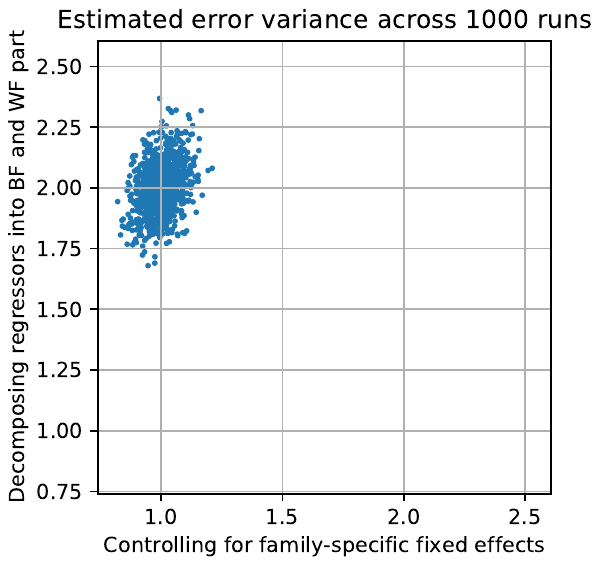 | **Code listing figure 3.** Estimated error variance based on OLS estimates of the model in Equation 1 (x-axis) and of the model in Equation 15 (y-axis), across 1000 runs in the simulation in **Code Listing 1**. |

**2–Supplemental Results**

**No indication of between-family sex different in genetic associations in the E-Risk**

We tested for sex differences in the E-Risk by estimating a cross-level interaction using a mixed effects model with a random intercept at the family level. Effect coding was used for Sex to facilitate interpretation. Thus, the intercepts represent the model grand mean of externalizing behavior across the categories in the model (i.e., male, female) for someone of average PGI. The main effects are interpretable as true main effects and not marginal effects. The interaction terms are directly interpretable as the difference in the association (i.e., slope) between the externalizing PGI and phenotypic externalizing for the effect group (i.e., males for both samples). Bias-adjusted 95% confidence intervals were produced using N=1000 sex-stratified bootstrapped samples to ensure stable interaction estimates. We did not observe any statistically significant between-pair sex differences (**Table S4** and **Figure S1A**), suggesting that the population and direct genetic associations of the externalizing PGI did not depend on the whether the twin pairs are comprised of all boys or all girls.

**3–Supplemental Figures**

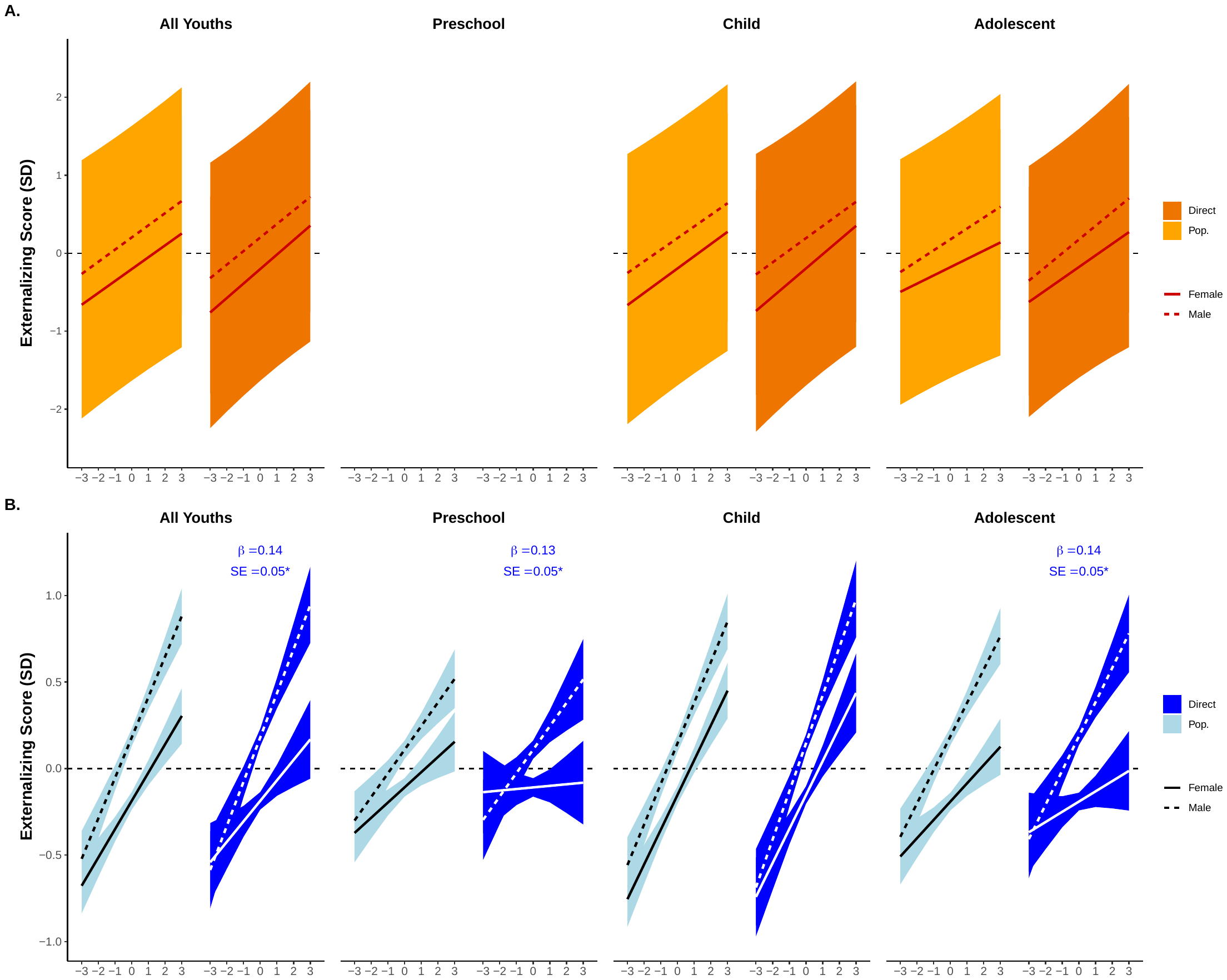

**Figure S1. Marginal effects of sex difference analysis**. **Panel A**. Between family sex differences in the E-Risk cohort estimated with a mixed linear model. **Panel B**. Sex differences in the MCS cohort estimated with a linear model.

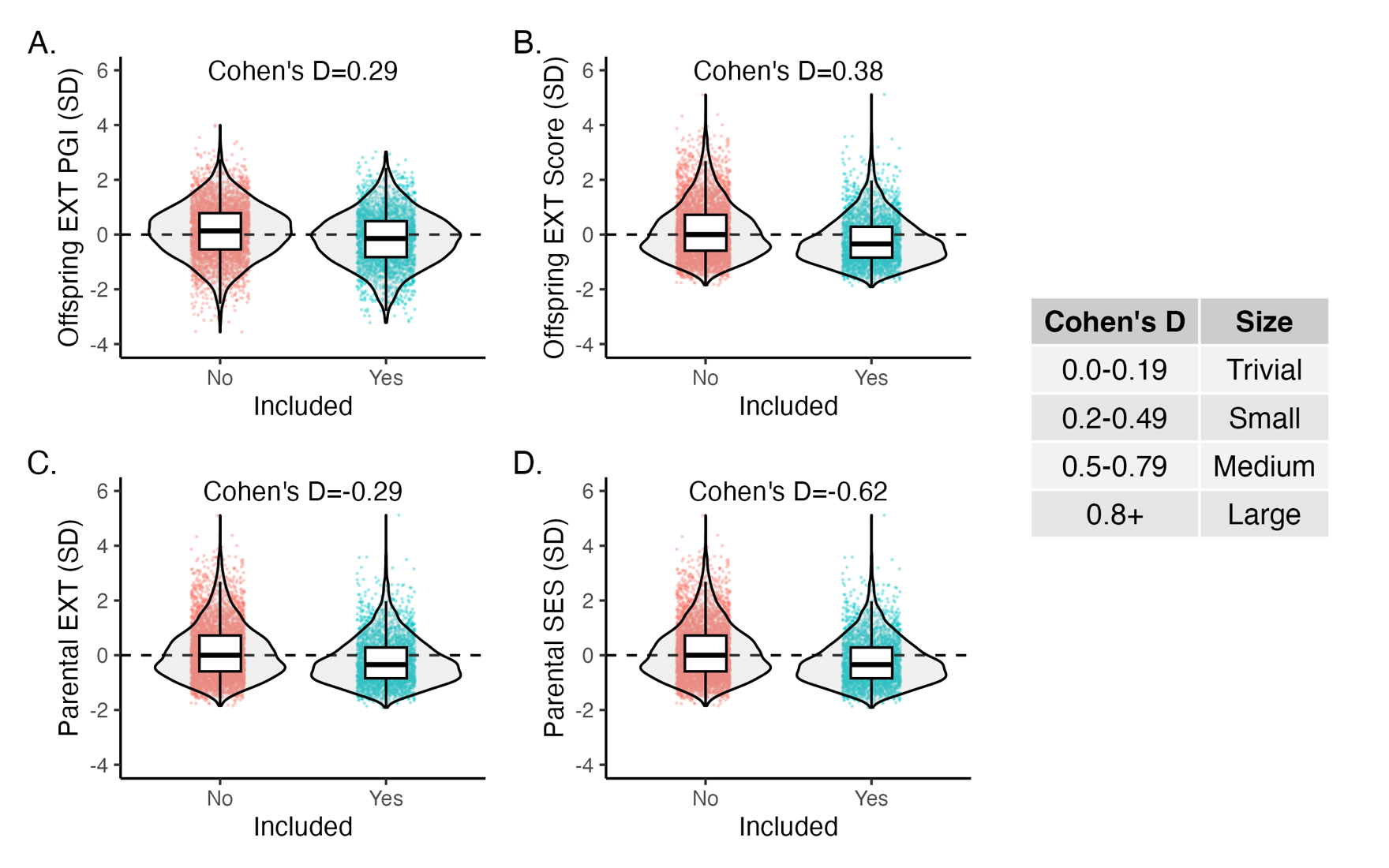

**Figure S2. Violin plots comparing distributional differences of key study variables across MCS participants who were and were not included in the final analytical sample due to data/family structure completeness.** Panels refer to main participant (offspring) PGI (**A**), participant (between-epoch) externalizing score (**B**), parental externalizing score (**C**), and parental socioeconomic status (**D**).

**3–Supplemental Tables**

| **Table S1**. Genotype-phenotype correlations in the E-Risk (sibling pairs) and MCS (mother/father) cohorts diagnosing the presence of assortative mating on externalizing genetics. | | | | | | |
| --- | --- | --- | --- | --- | --- | --- |
| **E-Risk** | | | | | | |
|  | | | | *r* | 95% CI |  |
| Sibling pair externalizing PGI | | | | 0.504 | [0.434, 0.575] |  |
| **MCS** | | | | | | |
| Mother | | Father | |  |  |  |
| Genotype | Phenotype | Genotype | Phenotype | *r* | 95% CI | *P*-value |
| X |  | X |  | 0.032 | [-0.004, 0.067] | 0.081 |
| X | X |  |  | 0.022 | [-0.014, 0.057] | 0.236 |
|  |  | X | X | 0.037 | [0.001, 0.072] | 0.043 |
|  | X |  | X | 0.044 | [0.008, 0.079] | 0.016 |
| Note: The E-Risk sibling pair correlating is an interclass correlation and tests for the presence of assortative mating on externalizing genetics by whether correlations differ from *r*=0.5 (i.e., confidence intervals do not include 0.5). In the MCS, the product of maternal (paternal) genotype-phenotype correlations and maternal-paternal phenotype correlation gives the expected correlation under the assumptions of phenotypic assortment (i.e., no assortment on externalizing genetics): *r_Expected_*=0.022 × 0.037 × 0.044 = 0.00003. Abbreviations: CI=confidence interval. | | | | | | |

| **Table S2**. Leave-one-parent-out analyses comparing the effect of parent-specific genotype adjustment on the population effect of externalizing in the MCS cohort. | | | | | | | | | | | | | |
| --- | --- | --- | --- | --- | --- | --- | --- | --- | --- | --- | --- | --- | --- |
|  |  | **All Youths** | | | **Preschool** | | | **Child** | | | **Adolescent** | | |
| Effect | Parent | β | SE | 95% CI | β | SE | 95% CI | β | SE | 95% CI | β | SE | 95% CI |
| Pop. | None | 0.206 | 0.019 | [0.168, 0.242] | 0.126 | 0.02 | [0.086, 0.166] | 0.207 | 0.017 | [0.174, 0.241] | 0.151 | 0.018 | [0.115, 0.186] |
| Partial | Father | 0.208 | 0.021 | [0.167, 0.248] | 0.108 | 0.025 | [0.06, 0.156] | 0.22 | 0.021 | [0.179, 0.262] | 0.147 | 0.021 | [0.105, 0.189] |
| Partial | Mother | 0.198 | 0.022 | [0.154, 0.24] | 0.119 | 0.024 | [0.072, 0.164] | 0.207 | 0.021 | [0.165, 0.248] | 0.14 | 0.02 | [0.101, 0.18] |
| Direct | Both | 0.197 | 0.027 | [0.146, 0.252] | 0.09 | 0.029* | [0.033, 0.147] | 0.227 | 0.026 | [0.177, 0.279] | 0.129 | 0.028 | [0.076, 0.184] |
|  |  | Parental control (%) | | | Parental control (%) | | | Parental control (%) | | | Parental control (%) | | |
|  | Father | 84.7% | | | 47.4% | | | 40.1% | | | 80.3% | | |
|  | Mother | 46.7% | | | 82.9% | | | 89.9% | | | 51% | | |
| *p<.01. Unless otherwise designated, all models are significant at *p*<.001. Parental control (%) refers to the percentage of attenuation observed between the population and direct genetic effects of the externalizing behavior PGI attributable to a single parent, and it is calculated as: $Parental control \left( \% \right)=((\left( partial direct genetic effect \right)-(direct genetic effect))/(\left( population genetic effect \right)-\left( direct genetic effect \right)))\times100$. Abbreviations: CI=confidence interval; Direct=direct genetic effect; Partial=partial direct genetic effect (i.e., adjusting for one parent); Pop.=population genetic effect; SE=standard error. | | | | | | | | | | | | | |

| **Table S3**. Family-level covariate adjustment of the population and direct genetic effect on externalizing in the E-Risk and MCS cohorts. | | | | | | | | | | | | | |
| --- | --- | --- | --- | --- | --- | --- | --- | --- | --- | --- | --- | --- | --- |
|  |  | **E-Risk** | | | | | | | | | | | |
|  |  | **All Youths** | | | **Preschool** | | | **Child** | | | **Adolescent** | | |
| Effect | Model | β | SE | 95% CI | β | SE | 95% CI | β | SE | 95% CI | β | SE | 95% CI |
| Pop. | Base | 0.169 | 0.034* | [0.104, 0.236] |  |  |  | 0.162 | 0.033*** | [0.098, 0.228] | 0.144 | 0.035*** | [0.077, 0.213] |
| Pop. | + Parental EXT | 0.098 | 0.03** | [0.038, 0.157] |  |  |  | 0.092 | 0.03** | [0.034, 0.151] | 0.086 | 0.032** | [0.023, 0.15] |
| Pop. | + Parental EXT & SES | 0.082 | 0.03** | [0.023, 0.142] |  |  |  | 0.08 | 0.03** | [0.021, 0.139] | 0.066 | 0.032* | [0.002, 0.129] |
| Direct | Base | 0.127 | 0.065 | [-0.002, 0.254] |  |  |  | 0.135 | 0.066* | [0.004, 0.264] | 0.077 | 0.065 | [-0.05, 0.205] |
| Direct | + Parental EXT | 0.127 | 0.06* | [0.009, 0.244] |  |  |  | 0.135 | 0.061* | [0.014, 0.254] | 0.077 | 0.062 | [-0.044, 0.199] |
| Direct | + Parental EXT & SES | 0.127 | 0.059* | [0.009, 0.242] |  |  |  | 0.135 | 0.061* | [0.014, 0.253] | 0.077 | 0.06 | [-0.041, 0.195] |
|  |  | **MCS** | | | | | | | | | | | |
|  |  | **All Youths** | | | **Preschool** | | | **Child** | | | **Adolescent** | | |
| Effect | Model | β | SE | 95% CI | β | SE | 95% CI | β | SE | 95% CI | β | SE | 95% CI |
| Pop. | Base | 0.206 | 0.019*** | [0.168, 0.242] | 0.126 | 0.02*** | [0.086, 0.166] | 0.207 | 0.017*** | [0.174, 0.241] | 0.151 | 0.018*** | [0.115, 0.186] |
| Pop. | + Parental EXT | 0.206 | 0.018*** | [0.171, 0.241] | 0.094 | 0.021*** | [0.055, 0.137] | 0.208 | 0.017*** | [0.174, 0.242] | 0.122 | 0.017*** | [0.09, 0.158] |
| Pop. | + Parental EXT & SES | 0.173 | 0.017*** | [0.14, 0.208] | 0.094 | 0.02*** | [0.055, 0.134] | 0.187 | 0.017*** | [0.152, 0.22] | 0.122 | 0.018*** | [0.087, 0.158] |
| Direct | Base | 0.197 | 0.027*** | [0.146, 0.252] | 0.09 | 0.029** | [0.033, 0.147] | 0.227 | 0.026*** | [0.177, 0.279] | 0.129 | 0.028*** | [0.076, 0.184] |
| Direct | + Parental EXT | 0.197 | 0.027*** | [0.143, 0.249] | 0.085 | 0.029** | [0.03, 0.144] | 0.228 | 0.027*** | [0.176, 0.281] | 0.13 | 0.026*** | [0.076, 0.18] |
| Direct | + Parental EXT & SES | 0.197 | 0.026*** | [0.145, 0.248] | 0.087 | 0.029** | [0.031, 0.143] | 0.228 | 0.027*** | [0.177, 0.277] | 0.13 | 0.026*** | [0.079, 0.181] |
| *p<.05, **p<.01, ***p<.001. Note: Model=individual nested models with increasing numbers of covariates. Abbreviations: CI=confidence interval; SE=standard error. | | | | | | | | | | | | | |

| **Table S4**. Sex differences in the population and direct genetic effects on externalizing in the E-Risk (cross-level interaction) and MCS (linear model) cohorts. | | | | | | | | | | | | | |
| --- | --- | --- | --- | --- | --- | --- | --- | --- | --- | --- | --- | --- | --- |
|  |  | **E-Risk** | | | | | | | | | | | |
|  |  | **All Youths** | | | **Preschool** | | | **Child** | | | **Adolescent** | | |
| Model | Coef. | β | SE | 95% CI | β | SE | 95% CI | β | SE | 95% CI | β | SE | 95% CI |
| Pop. | Intercept | 0.00 | 0.003 | [-0.006, 0.005] |  |  |  | 0.00 | 0.003 | [-0.005, 0.005] | 0.00 | 0.002 | [-0.005, 0.004] |
| Pop. | EXT_PGI_ | 0.154 | 0.024*** | [0.111, 0.206] |  |  |  | 0.153 | 0.024*** | [0.108, 0.203] | 0.123 | 0.024*** | [0.083, 0.176] |
| Pop. | Male | 0.406 | 0.033*** | [0.341, 0.47] |  |  |  | 0.39 | 0.035*** | [0.321, 0.458] | 0.356 | 0.034*** | [0.288, 0.421] |
| Pop. | EXT_PGI_ $\times$Male | 0.004 | 0.048 | [-0.093, 0.095] |  |  |  | -0.008 | 0.049 | [-0.109, 0.084] | 0.034 | 0.044 | [-0.053, 0.121] |
| Direct | Intercept | 0.00 | 0.002 | [-0.005, 0.005] |  |  |  | 0.00 | 0.003 | [-0.005, 0.005] | 0.00 | 0.002 | [-0.004, 0.003] |
| Direct | EXT_PGI_ | 0.124 | 0.042** | [0.042, 0.206] |  |  |  | 0.132 | 0.044** | [0.046, 0.217] | 0.076 | 0.04 | [-0.001, 0.155] |
| Direct | Male | 0.404 | 0.033*** | [0.339, 0.469] |  |  |  | 0.388 | 0.035*** | [0.32, 0.457] | 0.353 | 0.034*** | [0.285, 0.418] |
| Direct | EXT_PGI_ $\times$Male | 0.018 | 0.082 | [-0.142, 0.178] |  |  |  | 0.015 | 0.086 | [-0.154, 0.182] | 0.035 | 0.076 | [-0.112, 0.186] |
|  |  | **MCS** | | | | | | | | | | | |
|  |  | **All Youths** | | | **Preschool** | | | **Child** | | | **Adolescent** | | |
| Model | Coef. | β | SE | 95% CI | β | SE | 95% CI | β | SE | 95% CI | β | SE | 95% CI |
| Pop. | Intercept | -0.002 | 0.004 | [-0.011, 0.006] | -0.001 | 0.003 | [-0.007, 0.005] | -0.003 | 0.005 | [-0.012, 0.006] | -0.001 | 0.003 | [-0.008, 0.006] |
| Pop. | EXT_PGI_ | 0.202 | 0.017*** | [0.168, 0.235] | 0.117 | 0.018*** | [0.082, 0.154] | 0.219 | 0.017*** | [0.186, 0.253] | 0.15 | 0.018*** | [0.115, 0.185] |
| Pop. | Male | 0.357 | 0.034*** | [0.29, 0.424] | 0.216 | 0.037*** | [0.143, 0.288] | 0.292 | 0.035*** | [0.222, 0.359] | 0.369 | 0.034*** | [0.304, 0.439] |
| Pop. | EXT_PGI_ $\times$Male | 0.063 | 0.033 | [-0.003, 0.127] | 0.044 | 0.038 | [-0.031, 0.118] | 0.028 | 0.036 | [-0.042, 0.099] | 0.082 | 0.036 | [0.01, 0.151] |
| Direct | Intercept | -0.002 | 0.005 | [-0.011, 0.007] | 0.00 | 0.003 | [-0.007, 0.006] | -0.003 | 0.005 | [-0.013, 0.006] | -0.001 | 0.004 | [-0.008, 0.006] |
| Direct | EXT_PGI_ | 0.192 | 0.026*** | [0.14, 0.244] | 0.081 | 0.028** | [0.028, 0.139] | 0.239 | 0.028*** | [0.186, 0.295] | 0.128 | 0.026*** | [0.077, 0.18] |
| Direct | Male | 0.357 | 0.033*** | [0.293, 0.424] | 0.215 | 0.039*** | [0.138, 0.29] | 0.293 | 0.034*** | [0.226, 0.361] | 0.369 | 0.035*** | [0.3, 0.439] |
| Direct | EXT_PGI_ $\times$Male | 0.123 | 0.051* | [0.024, 0.223] | 0.115 | 0.054* | [0.007, 0.219] | 0.069 | 0.056 | [-0.04, 0.181] | 0.132 | 0.054* | [0.026, 0.24] |
| *p<.05, **p<.01, ***p<.001. Note: Effect coding (i.e., not dummy coding) was used for both analyses. Only between-pair sex differences could be estimated in the E-Risk cohort because twins were all same-sex pairs. Between family sex differences were estimated as a cross-level interaction between EXT_PGI_ and Sex in a mixed linear model, with a random intercept at the family level. Sex differences in the MCS were estimated using a linear model. Abbreviations: CI=confidence interval; SE=standard error. | | | | | | | | | | | | | |

| **Table S5**. Comparison between MCS participants who were and were not included in the final analytical sample on key study variables. | | | | |
| --- | --- | --- | --- | --- |
|  |  | Included in Analytic Sample | |  |
|  | Overall (N=8201) | No (N=5377) | Yes (N=2824) | P-value* |
| Sex |  |  |  |  |
| Female | 4093 (49.9%) | 2693 (50.1%) | 1400 (49.6%) | .679 |
| Male | 4108 (50.1%) | 2684 (49.9%) | 1424 (50.4%) |  |
| EXT Score (z-score) |  |  |  |  |
| Mean (SD) | 0.00 (1.00) | 0.135 (1.04) | -0.237 (0.887) | <.001 |
| Missing | 433 (5.3%) | 433 (8.1%) | 0 (0%) |  |
| EXT PGI (z-score) |  |  |  |  |
| Mean (SD) | 0.00 (1.00) | 0.118 (0.996) | -0.166 (0.982) | <.001 |
| Missing | 1403 (17.1%) | 1403 (26.1%) | 0 (0%) |  |
| Parental EXT (z-score) |  |  |  |  |
| Mean (SD) | 0.00 (1.00) | -0.110 (1.09) | 0.173 (0.801) | <.001 |
| Missing | 915 (11.2%) | 915 (17.0%) | 0 (0%) |  |
| Parental SES (z-score) |  |  |  |  |
| Mean (SD) | 0.00 (1.00) | -0.206 (1.03) | 0.393 (0.803) | <.001 |
| Missing | 2 (0.0%) | 2 (0.0%) | 0 (0%) |  |
| *P-value refers to chi-square test for categorical variables and t-test for continuous variables. Comparisons were between the MCS participants who were and were not included in the final analytical sample. | | | | |

**4–Supplemental References**

(7) Martin, A. R.; Gignoux, C. R.; Walters, R. K.; Wojcik, G. L.; Neale, B. M.; Gravel, S.; Kenny, E. E. Human demographic history impacts genetic risk prediction

across diverse populations. *American Journal of Human Genetics* **2017**, *100*, 635–649.

(23) Achenbach, T. M.; Edelbrock, C. S. Manual for the Child: Behavior Checklist and Revised Child Behavior Profile. 1983.

(24) Goodman, R. The Strengths and Difficulties Questionnaire: a research note. *J Child Psychol Psychiatry* **1997**, *38* (5), 581-586. DOI: 10.1111/j.1469-7610.1997.tb01545.x.

(25) Costa, P. T.; McCrae, R. R. Normal personality assessment in clinical practice: The NEO Personality Inventory. *Psychological assessment* **1992**, *4* (1), 5.
